# Impacts of the COVID-19 pandemic on antibiotic use and resistance in hospitals: a retrospective ecological analysis of French national surveillance data over 2019-2022

**DOI:** 10.1101/2024.12.04.24317990

**Authors:** Maylis Layan, David R M Smith, Solen Kernéis, Loïc Simon, Catherine Dumartin, Lory Dugravot, Amélie Jouzeau, Sylvie Maugat, Laetitia Gambotti, Laurence Watier, Lulla Opatowski, Laura Temime

## Abstract

**Summary:** *Background:* The COVID-19 pandemic led to major disruptions in healthcare services at the hospital and community levels. The resulting impact on antibiotic resistance (ABR) in hospitals is difficult to predict.

*Methods:* We exploited data from the French national surveillance system over four years (2019-2022) including 414 hospitals across 12 French regions. We evaluated changes in annual antibiotic use compared to 2019 using multiple comparison tests. We also compiled a large dataset of 692,551 incident isolates for five antibiotic-bacterium pairs. Using negative binomial regression models accounting for autocorrelation and antibiotic use, we evaluated associations between resistant isolates incidence and COVID-19 indicators (pandemic periods or intubated COVID-19 patient prevalence). We investigated how these associations varied specifically in ICUs (*n*=85) and across geographical regions.

*Findings:* The use of some antibiotics, including azithromycin, imipenem and meropenem, significantly increased between 2020 and 2022. Concomitantly, the incidence of methicillin-resistant *Staphylococcus aureus* (up to 37%, 95% CI: 18-53%) and ESBL-producing *Escherichia coli* (up to 33%, 95% CI: 16-46%) isolates significantly decreased in hospitals and ICUs during the pandemic. A transient decrease in ICUs was also observed for ESBL-producing *Klebsiella pneumoniae* during periods of strong anti-COVID-19 interventions in the community (24%, 95% CI: 6-38%). No significant changes for ESBL-producing *Enterobacter cloacae* complex were observed. Very interestingly, the incidence of carbapenem-resistant *Pseudomonas aeruginosa* isolates was associated with COVID-19 intubation prevalence in hospitals (p<0.001) and ICUs (p<0.001), notably in the regions most affected by the pandemic.

*Interpretation:* Our results highlight strong modifications of antibiotic use and pathogen-specific global impacts of the COVID-19 pandemic on ABR in hospitals. Even though the biological mechanisms underlying between-species differences remain unclear, these results provide important insights into the potential impacts of a viral pandemic on ABR and support the need for pandemic preparedness in healthcare facilities.

*Funding:* ANR-10-LABX-62-IBEID.

*Research in context:* Evidence before this study
We searched PubMed for articles in English published between Jan 1, 2020, and August 31, 2024 exploring national-scale changes in antibiotic resistance (ABR) within healthcare settings during the COVID-19 pandemic. Search terms for titles and abstracts were (“antibiotic resistance” OR “antimicrobial resistance” OR “bacterial resistance” OR “drug resistance” OR “MRSA” OR “ESBL” OR “carbapenem resistant”) AND (“hospital” OR “healthcare”) AND (“COVID-19” OR “SARS-CoV-2”) AND (“national” OR “nationwide”). The search yielded 94 results. We identified 12 relevant studies after filtering out articles referring to viruses, parasites, or fungi, focusing on a single hospital, evaluating changes in antibiotic use only, assessing healthcare workers’ practices, or using qualitative approaches. All studies used either national surveillance data on antibiotic resistance or large multi-center cohorts of inpatients. Five studies showed a significant increase in MRSA, at least during the first wave of the COVID-19 pandemic, while two studies did not find changes at the national level compared to 2019. Interestingly, one study showed that the abandonment of infection prevention and control strategies that specifically target hospital-acquired MRSA infections was associated with an increase of these infections, regardless of COVID-19 admission prevalence. One study in Spain showed decreased resistance of *P. aeruginosa* to all tested antibiotics in 2022 compared to 2017, using point prevalence survey results. Conversely, one study focusing on US Veterans Affairs hospitals showed increased incidence and resistance of healthcare-associated carbapenem-resistant *P. aeruginosa* (CR-PA) infections during the pandemic. Most studies used univariate statistical approaches. Only two studies included COVID-19-related variables in their models; they found no association with MRSA or extended-spectrum cephalosporin-resistant *E. coli* and *K. pneumoniae*. Added value of this study
Here, we provide the first evaluation of the impact of the pandemic on antibiotic consumption and resistance for five antibiotic-bacterium pairs (MRSA, CR-PA, ESBL-producing *E. coli*, ESBL-producing *K. pneumoniae*, and ESBL-producing *E. cloacae* complex) in hospitals, at the national and regional scales. By analyzing French surveillance data from the SPARES database including 414 hospitals that represent up to 14% of French hospitals, we evaluated annual changes in antibiotic use and quantified the impacts at the weekly level of the COVID-19 pandemic on the incidence of five of the most prevalent resistant bacteria in France. Accounting for autocorrelation and antibiotic use, factors that were not considered in previous studies, we report a significant positive association between the weekly incidence of CR-PA isolates and the prevalence of intubated COVID-19 patients in the preceding weeks. Carbapenem use and intubation being risk factors of CR-PA infections, our results suggest a direct impact of the pandemic on CR-PA epidemiology. Inversely, we show that the incidence of ESBL-producing *E. coli* and MRSA isolates decreased after the start of the first pandemic wave at the hospital level but also in ICUs. The fine grain analysis across 12 French administrative regions revealed regional heterogeneities, but highlighted consistent associations in the regions most affected by the COVID-19 pandemic. Implications of all the available evidence
Pandemics not only destabilize healthcare systems by adding pressure and changing healthcare worker behaviors, but also influence the epidemiology of other infectious diseases as shown in our study. We specifically highlight the contrasting effects of the COVID-19 pandemic on ABR in French hospitals, associated with an increase in CR-PA isolate incidence but a general decrease in ESBL-producing *E. coli* and MRSA. This work highlights how national-scale hospital surveillance systems such as SPARES that collect data at the weekly level are key to capture the evolving impacts of pandemics. They also allow to generate hypotheses on the potential mechanisms of action of the pandemic on ABR epidemiology, as showcased by the analysis of CR-PA isolates incidence, and thereby participate in the improvement of healthcare systems in pandemic context.

## Introduction

Antibiotic resistance (ABR) is a leading health problem worldwide associated with an estimated 1.14 (1.00- 1.28) million deaths in 2021.^1^ Exceptional health events, such as pandemics, lead to major changes in care and hygiene practices in both the community and hospitals, which may modify ABR epidemiology. For instance, during the COVID-19 pandemic, social distancing measures^2^ and reduced outpatient antibiotic prescription^3,4^ might have reduced ABR burden in hospitals. Healthcare workers (HCWs) also reported greater hand hygiene compliance and availability of alcohol-based hand rub.^5,6^ On the other hand, inpatient antibiotic use increased in France,^7^ probably related to the high proportion of COVID-19 patients receiving antibiotics as reported in several meta-analyses.^8,9^ In parallel, surges in COVID-19 patients in hospitals often exceeded bed capacity with a sicker patient-case mix.^5^ This generated higher workload and decreased the time that HCWs could allocate to antibiotic stewardship and infection prevention and control (IPC).^5^ A striking example is the increase of MRSA incidence in US Veterans Affairs hospitals with interrupted IPC during the pandemic.^10^ The combined impact of all these effects, at the community and hospital levels, is difficult to predict^11^ and may have changed throughout the pandemic, which was notably marked by the improvement of COVID-19 patient management and treatment (e.g. reduction in antibiotic prescribing^12,13^ and mechanical ventilation^14^).

Several studies have attempted to quantify the impacts of the pandemic on ABR in hospitals, but the evidence remains conflicting.^10,15,16^ A meta-analysis estimated that MRSA incidence did not change during the pandemic, whereas there was a statistically insignificant trend for increased incidence of extended-spectrum β-lactamase (ESBL)-producing Enterobacterales and CR-PA.^15^ Heterogeneity in terms of settings, study design, and health outcomes, as well as heterogeneity in baseline epidemiological situations and local practices, might have undermined the statistical power of this meta-regression. A recent comprehensive study on antibiotic-resistant healthcare-associated infections in US Veterans Affairs hospitals also highlighted varying trends across antibiotic-bacterium combinations during the pandemic.^16^

Here, we aimed to assess the impacts of the pandemic on antibiotic use and antibiotic-resistant bacteria in French hospitals using large-scale surveillance data over 2019-2022. We evaluated changes in antibiotic use in hospitals compared to 2019 as a potential key mechanism of pandemic action on ABR epidemiology in hospitals. For five antibiotic-resistant bacteria of major importance in French hospitals (ESBL-producing *E. coli* - ESBL-EC, ESBL-producing *K. pneumoniae* - ESBL-KP, ESBL-producing *E. cloacae* complex - ESBL-ECC, MRSA, and CR-PA), we quantified the association between their incidence and the burden of severe COVID-19 patients, we investigated whether these associations were similar in the specific context of intensive care units (ICUs), and we evaluated how associations with the pandemic varied across geographical regions.

## Methods

### Data sources

#### Antibiotic resistance and antibiotic consumption in hospitals

We obtained ABR and antibiotic use data from the French National Surveillance Database on Antibiotic Resistance in hospitals (SPARES). SPARES compiles data reported by participating hospitals on (*i*) microbiological test results from clinical samples and (*ii*) annual antibiotic consumption by anatomical therapeutic chemicals (ATC) class (Figure 1A).^7^ Consolidated data are only available from 2019 on. SPARES provided us with microbiological test results and antibiotic use data from the 1^st^ of January 2019 to the 31^st^ of December 2022.

**Figure 1.**
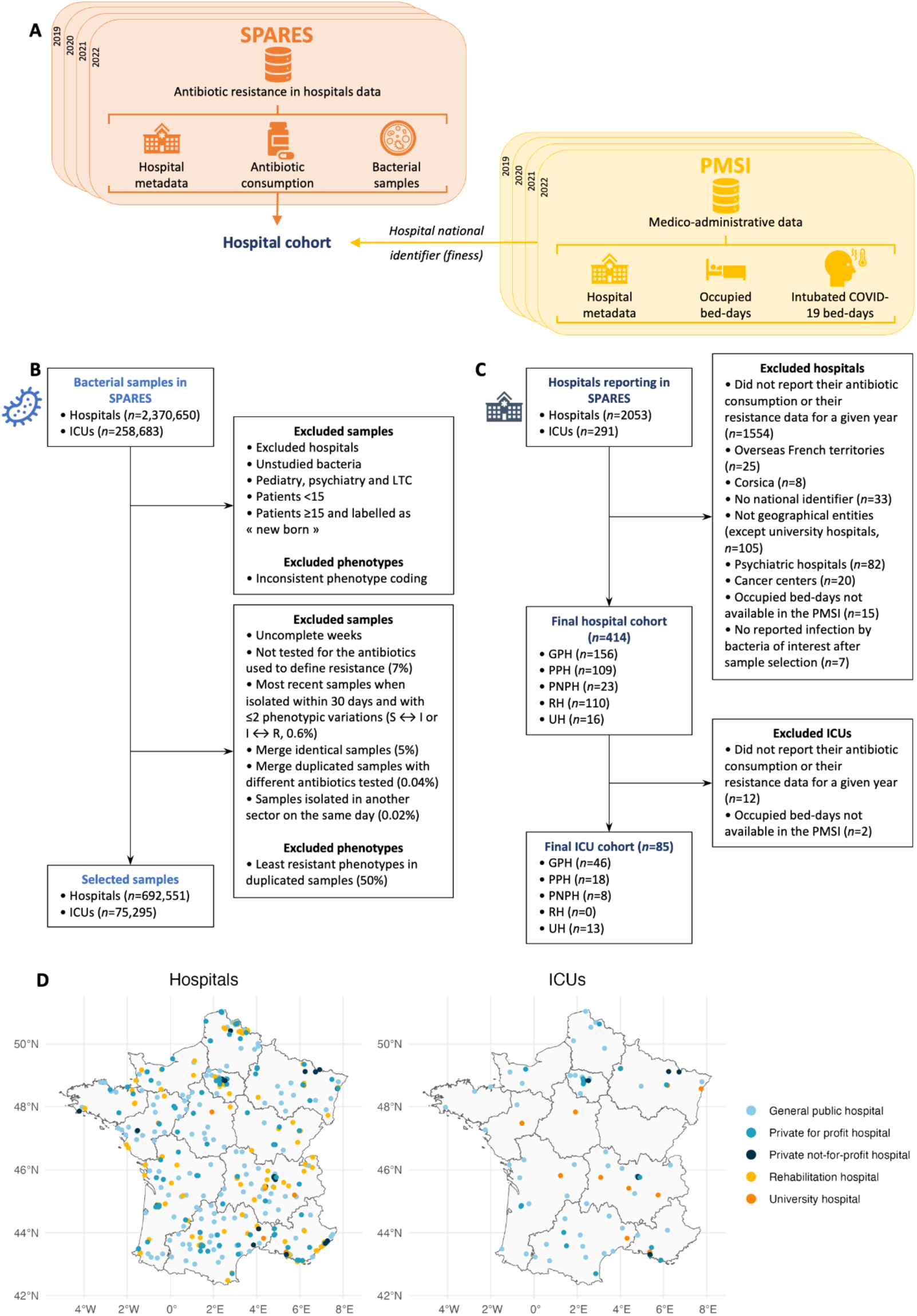
Data sources and selection. (**A**) Hospitals report their antibiotic resistance data in SPARES using either their geographical identifier or their legal identifier, the latter generally grouping multiple geographical entities. We excluded all hospitals that use their legal identifier, except for teaching hospitals that often host the most severe cases. For teaching hospitals that report their data under their legal identifier, we retrieved occupied bed-days data from the PMSI using the geographical identifiers of all relevant entities (see appendix pp 3-6 for more details). (**B**) Flow diagram of sample selection. We focused our investigations on meticillin-resistant *S. aureus*, ESBL-producing *E. coli*, *K. pneumoniae*, and *E. cloacae* complex, and carbapenem-resistant *P. aeruginosa*. *E. cloacae* complex include *Enterobacter cloacae*, *Enterobacter absuriae*, *Enterobacter hormaechei*, *Enterobacter kobei*, *Enterobacter ludwigii*, and *Enterobacter nimipressuralis*. Among the duplicated samples with different phenotypes, we excluded the least resistant phenotypes, meaning phenotypes S if R or I, and phenotypes I if R (2,219/4,442, 50%). In total, we selected 34% of isolates from the global database and 36% of samples isolated in intensive care units (ICUs). (**C**) Flow diagram of hospitals and ICUs selection. Hospitals located in French overseas territories and Corsica were removed due to specific epidemiological situations as well as low numbers that would decrease the statistical power of the regional analyses. (**D**) Geographical distribution of hospitals and ICUs across the 12 regions of mainland France. GPH: general public hospital; PPH: private for profit hospital; PNPH: private not-for-profit hospital; RH: rehabilitation hospital; UH: university hospital.

Clinical tests reported in SPARES encompass infection and colonization events. No information about the source of acquisition (community or healthcare) is available. Clinical test results are interpreted according to the EUCAST guidelines by participating hospitals.

Hospitals report their annual antibiotic consumption by ATC class in number of defined daily doses (DDD) for 1,000 bed-days.

#### Hospital stays

We obtained data on occupied bed-days from the National Hospital Discharge Database (PMSI), a database dedicated to hospital activity evaluation for budget allocation. As such, it documents for all hospital stays their type, duration, associated diagnoses and medical procedures. We exploited this database to extract the weekly number of occupied bed-days in acute care facilities between the 1^st^ of January 2019 and the 31^st^ of December 2022. We also extracted the weekly number of intubated COVID- 19 patients’ bed-days (appendix pp 6-7).

### Data selection procedure

We explored ESBL-EC, ESBL-ECC, ESBL-KP, MRSA, and CR-PA dynamics since they are on the WHO priority list^17^ and amongst the resistant bacteria with highest incidence in France.^18^ Of all available isolates in SPARES (*n*=2,370,650), we selected the ones corresponding to the studied bacterial species that are tested for the resistance of interest. Importantly, when multiple samples potentially corresponded to the same acquisition episode and were performed within a 30-day period, we kept only the first sample as we were interested in incident episodes (Figure 1B). Our selection procedure led to the inclusion of 692,551 episodes, 74,387 of which were resistant. Concerning antibiotics, only ATC-J01 antibiotics reported over the four years of the study period were included (appendix pp 2-3).

Hospitals report their data to SPARES on a voluntary basis leading to non-exhaustive and possibly non representative data collection. To ensure our analysis is based on a temporally stable database, we restricted our analyses to hospitals that systematically reported their antibiotic consumption and clinical tests results over the four years of the study period. Other exclusion criteria are listed in Figure 1C. Selected hospitals (*n*=414) are distributed over the 12 administrative regions of hexagonal France (Figure 1D). We assessed the representativeness of our cohort in appendix pp 3-6.

### Outcomes

We investigated the dynamics of the following outcomes at the national and/or regional levels and at the hospital and/or ICU levels over the study period:

- Annual proportion of resistant episodes for each antibiotic-bacterium pair: annual number of resistant episodes divided by annual number of total episodes.
- Weekly incidence of resistant bacterial isolates: weekly number of incident resistant episodes divided by weekly number of occupied bed-days (from the PMSI) times 1,000.
- Annual consumption of antibiotics in DDD for 1,000 bed-days.

The hospital level included rehabilitation care, general medicine, surgery, gynecology-obstetrics, and ICU.

### Statistical analyses

#### Antibiotic consumption dynamics

For each antibiotic class, we evaluated whether its consumption had changed over the study period by comparing annual consumption distributions using Friedman tests (non-parametric alternative of the one-way ANOVA for repeated measures). We then explored the evolution of antibiotic consumption using 2019 as a reference year by performing three pairwise Wilcoxon signed-rank tests. We applied a Bonferroni correction for multiple testing, and reported adjusted p-values and 98.3% confidence intervals (CIs) to account for the correction.

#### Antibiotic-resistant episodes dynamics

For each antibiotic-bacterium pair, we evaluated whether there was a linear trend in the annual proportion of resistance in hospitals at the national level using the χ^2^ test for trend in proportions. When resistance proportions exhibited a significant increase or decrease between 2019 and 2022, we estimated the slope of the linear trend using linear regression minimizing weighted least squares.

Then, for each bacterium pair, we investigated ABR and COVID-19 association using multivariate count regression models. We modeled the incident number of resistant isolates of a bacterial species *N*_*w*_ during week *w* in hospitals or ICUs as a negative binomial distribution to account for overdispersion. The equations of the models are as follows:

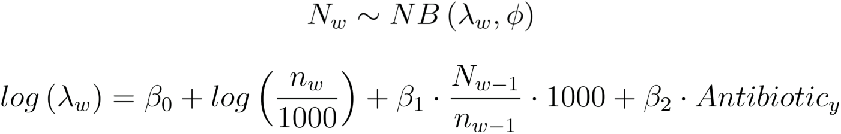

With *ϕ* the overdispersion parameter, *β*_0_ the intercept, *n*_*w*_ the number of occupied bed-days at week *w*, 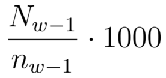 the occupied bed-days permillage at week *w* (offset),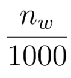 the incidence of resistant bacterial isolates per 1,000 bed-days in the preceding week (*w*−1) to account for autocorrelation, and *Antibiotic*_*y*_ the consumption level (in DDD for 1,000 bed-days) of the main antibiotic class targeted by the resistance during year *y* that includes week *w*. Target antibiotic classes correspond to imipenem+meropenem for *P. aeruginosa*, 3GC for Enterobacterales, and penicillins for *S. aureus*. We considered antibiotic consumption as a confusion factor, and investigated its impact in a sensitivity analysis (appendix pp 22-25). We compared this baseline model to 6 distinct models accounting for COVID-19-related variables (appendix p10-11). These variables were either the period of the pandemic (a categorical variable indicating the pre-pandemic period, the first wave and periods marked by three levels of anti-COVID-19 restrictions in the community), or COVID-19 intubation prevalence. The levels of anti-COVID-19 restrictions in the community were based on Paireau and colleagues,^2^ but we considered the first wave separately as it was marked by a very specific context of care disorganization in hospitals. The prevalence of intubated COVID-19 patients (weekly number of intubated COVID-19 patients’ bed-days divided by the weekly number of bed-days times 1,000) represented the burden of severe COVID-19 patients. For both COVID-19-related variables, we tested variables at week *w*, *w* − 1, and *w* − 2. Continuous explanatory variables (prevalence of COVID-19 intubation, antibiotic consumption, resistant isolate incidence at week *w* − 1) were on very different scales, so we standardized them to facilitate the comparison of regression coefficients across explanatory variables, models and bacterial species.

We calculated the 95% CIs for the regression coefficients using a normal approximation to the distribution of the maximum likelihood estimators. For each antibiotic-bacterium pair, we selected the model with the lowest Akaike Information Criterion (AIC). Models were fitted by maximum likelihood using the glm.nb function of the R package MASS (appendix pp 10-19).^19^

When the best model included COVID-19-related variables, we further explored how associations varied across the 12 administrative regions by fitting the model selected in the national analysis to regional hospital data separately for each region and accounting for overdispersion when necessary. We did not investigate regional ICU data alone due to the limited number of episodes and absence of ICUs in some regions (Figure 1D).

All statistical analyses were done with R, version 4.3.0. This study adheres to the Strengthening the Reporting of Observational studies in Epidemiology (STROBE) reporting guideline.

## Results

### Evolution of antibiotic use in hospitals during the pandemic in France

Annual antibiotic consumption strongly varied across hospitals (*n*=414, gray lines) of our cohort and across antibiotic classes, penicillins and quinolones being the most used (appendix p20 and Table 1). Overall, a median increase in total consumption of 7.5 DDD per 1,000 bed-days (98.3% CI: 2.7, 12.5) was found in 2020 compared to 2019 but it did not persist. For all antibiotic classes explored, significant changes in annual consumption were found, with strong variations across classes (Table 1). Carbapenem consumption, and more specifically imipenem and meropenem, showed a significant increase in 2020 and 2021, while cephalosporins increased until 2022. Interestingly, macrolide consumption significantly increased only in 2020 (+1.71 DDD/1,000 bed-days, 98.3% CI: +0.9 to +2.6), while azithromycin consumption significantly increased during the whole pandemic period.

**Table 1.**
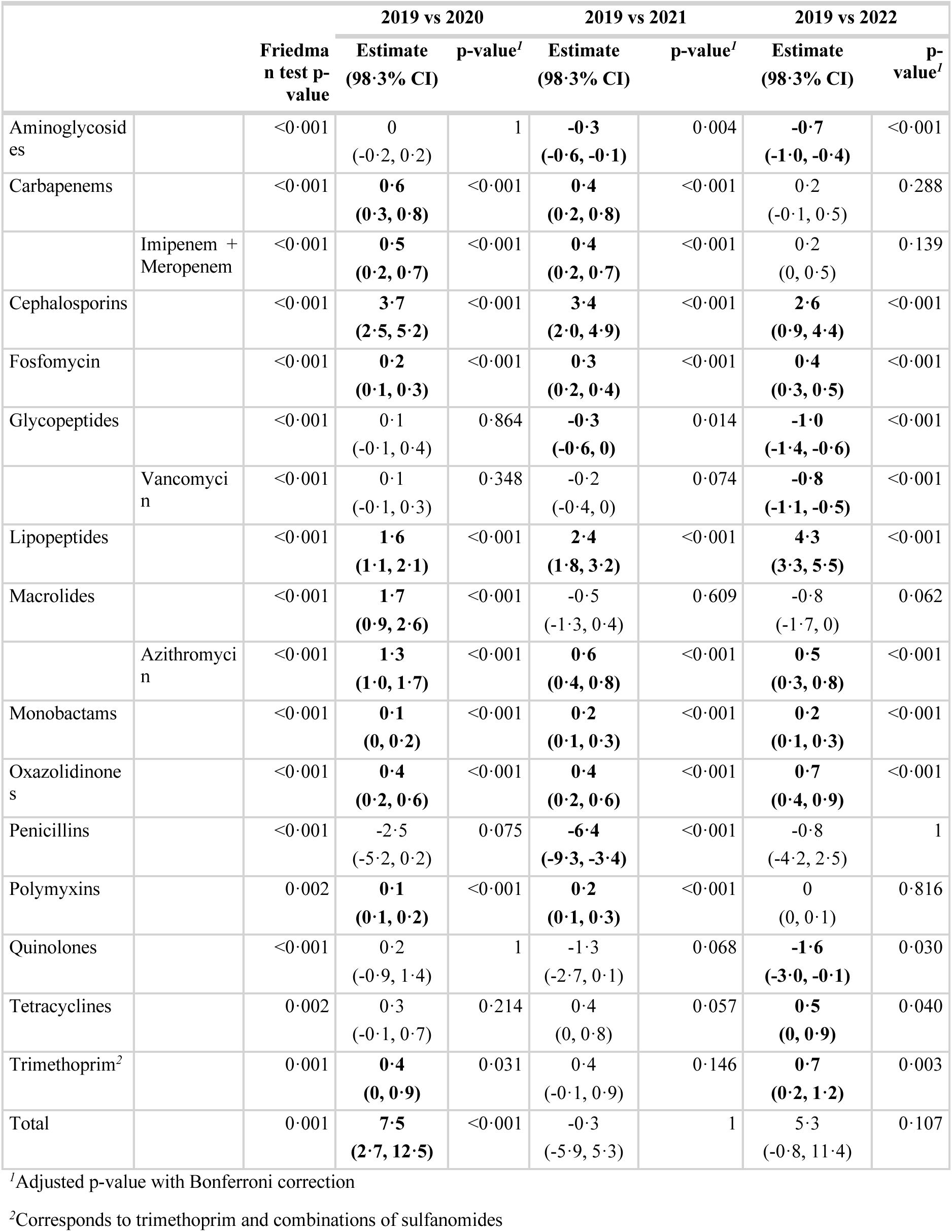
Changes in annual antibiotic consumption in French hospitals, 2019-2022. We report absolute differences in consumption at the hospital level for every antibiotic class, as well as imipenem and meropenem, vancomycin, and azithromycin specifically. Estimates correspond to the median of the differences of antibiotic consumption (in DDD per 1,000 bed-days) between two years of the study period, using 2019 as a reference year. Positive values thereby indicate an increased consumption compared to 2019 and negative values indicate a reduced consumption compared to 2019. For every pairwise comparison, we report the adjusted p-value of the paired Wilcoxon signed-rank test using a Bonferroni correction and the associated 98.3% CI. Finally, we report Friedman tests p-values that are the non-parametric equivalent of one-way repeated measures ANOVA tests to evaluate whether there are changes in antibiotic use over the study period.

Antibiotic use dynamics varied across regions (Figure 2A and appendix p 21). A significant increase of total antibiotic use was observed in 2020 in Île-de-France (IDF), Grand-Est (GES) and Auvergne-Rhône-Alpes (ARA). Interestingly, macrolide consumption significantly increased in four regions in 2020, especially azithromycin whose consumption increased in seven regions (Figure 2B). Importantly, the increase of azithromycin use persisted until 2022 in Hauts-de-France (HDF), Occitanie (OCC), and Provence-Alpes- Côte d’Azur (PAC).

**Figure 2.**
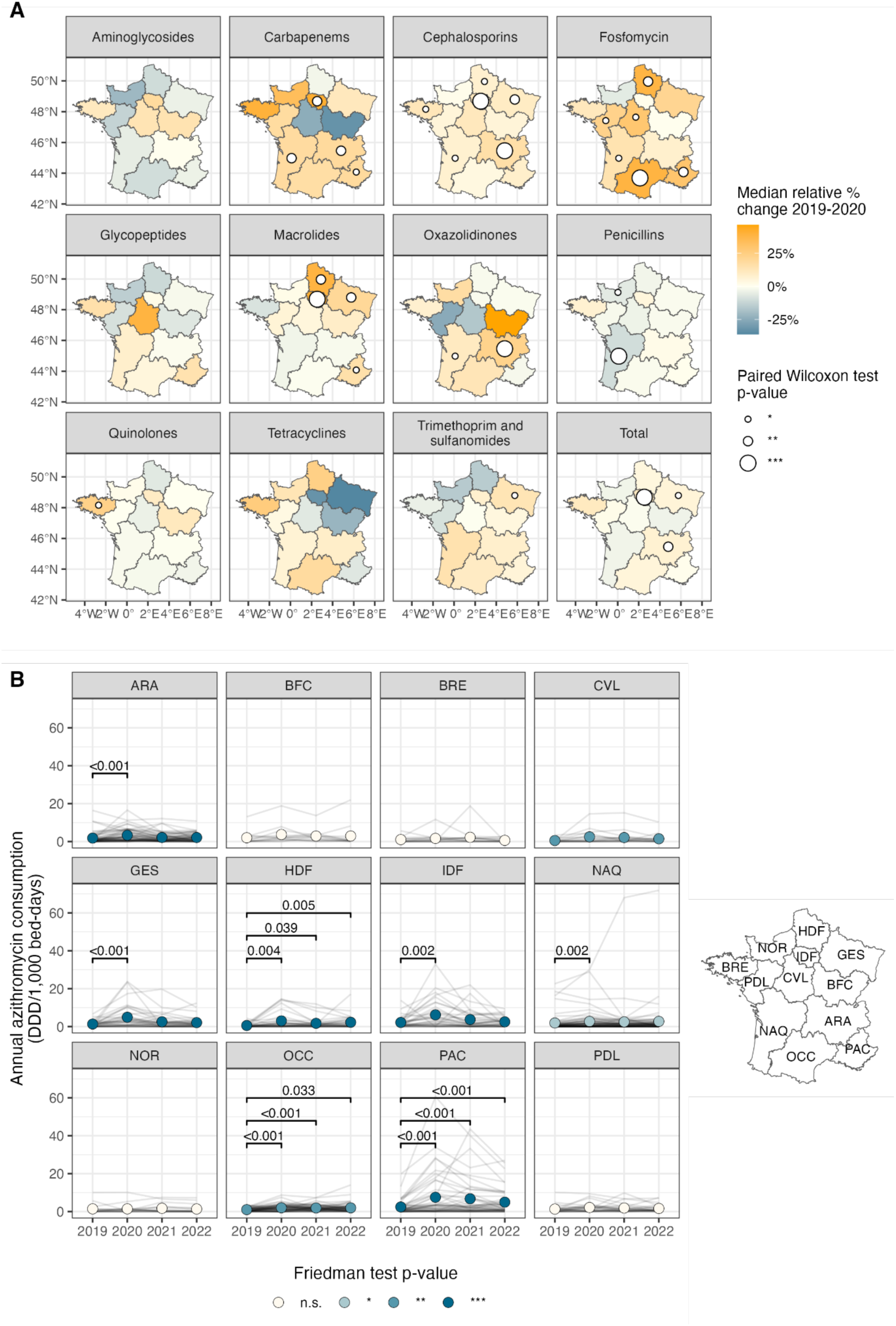
Regional heterogeneity in antibiotic consumption in French hospitals, 2019-2022. (**A**) Changes of antibiotic consumption across French regions between 2019 and 2020. Colors indicate the median percentage change of antibiotic consumption between 2019 and 2020. We excluded hospitals reporting no consumption in 2019 to calculate this metric. Dark blue colors indicate increased consumption and dark orange colors indicate decreased consumption. We also performed paired Wilcoxon signed-rank tests to compare regional distributions of antibiotic consumption in 2019 and 2020. Circle size indicates the level of significance of the paired Wilcoxon signed-rank tests when the p-value is ≤0.05. (**B**) Dynamics of azithromycin consumption across French regions during the study period. Each panel represents a French region whose geographical location is shown on the right. Gray lines correspond to the trajectories of individual hospitals and colored circles to the average regional antibiotic consumption in defined daily doses (DDD) for 1,000 occupied bed-days. Colors indicate the level of significance of the Friedman test which is the non-parametric equivalent of one-way repeated measures ANOVA tests. We also indicate the corrected p-values of the paired Wilcoxon signed-rank tests between 2019 and the other years of the study period when corrected p- values are ≤0.05. We used a Bonferroni correction. n.s. : p-value>0.05; *: p-value≤0.05; **: p-value≤0.01; ***: p-value≤0.001. ARA: Auvergne-Rhône-Alpes; BFC: Bourgogne-Franche-Comté; BRE: Bretagne; CVL: Centre-Val de Loire; GES: Grand-Est; HDF: Hauts-de-France; IDF: Île-de-France ; NAQ: Nouvelle-Aquitaine; NOR: Normandie; OCC: Occitanie; PAC: Provence-Alpes-Côte d’Azur; PDL: Pays de la Loire.

In ICUs (*n*=85), we observed higher antibiotic consumption levels (appendix p20) and changes of larger magnitude compared to the overall hospital level (Table 2). Although we did not find a significant change in total antibiotic consumption in these wards, we observed a significantly increased use of carbapenems, cephalosporins, and macrolides in 2020. Imipenem and meropenem consumption remained significantly higher until 2022 (+12.42 DDD/1,000 bed-days, 98.3% CI: +1.9 to +22.4), while carbapenem use decreased to its 2019 level in 2022. Macrolide (+17.41 DDD/1,000 bed-days, 98.3% CI: +7.7 to +28.9) and azithromycin (+12.69 DDD/1,000 bed-days, 98.3% CI: +5.8 to +20.5) consumption significantly increased but only during the first year of the pandemic, contrary to the hospital level. Finally, penicillin consumption significantly decreased in 2020 and 2021 compared to 2019.

**Table 2.**
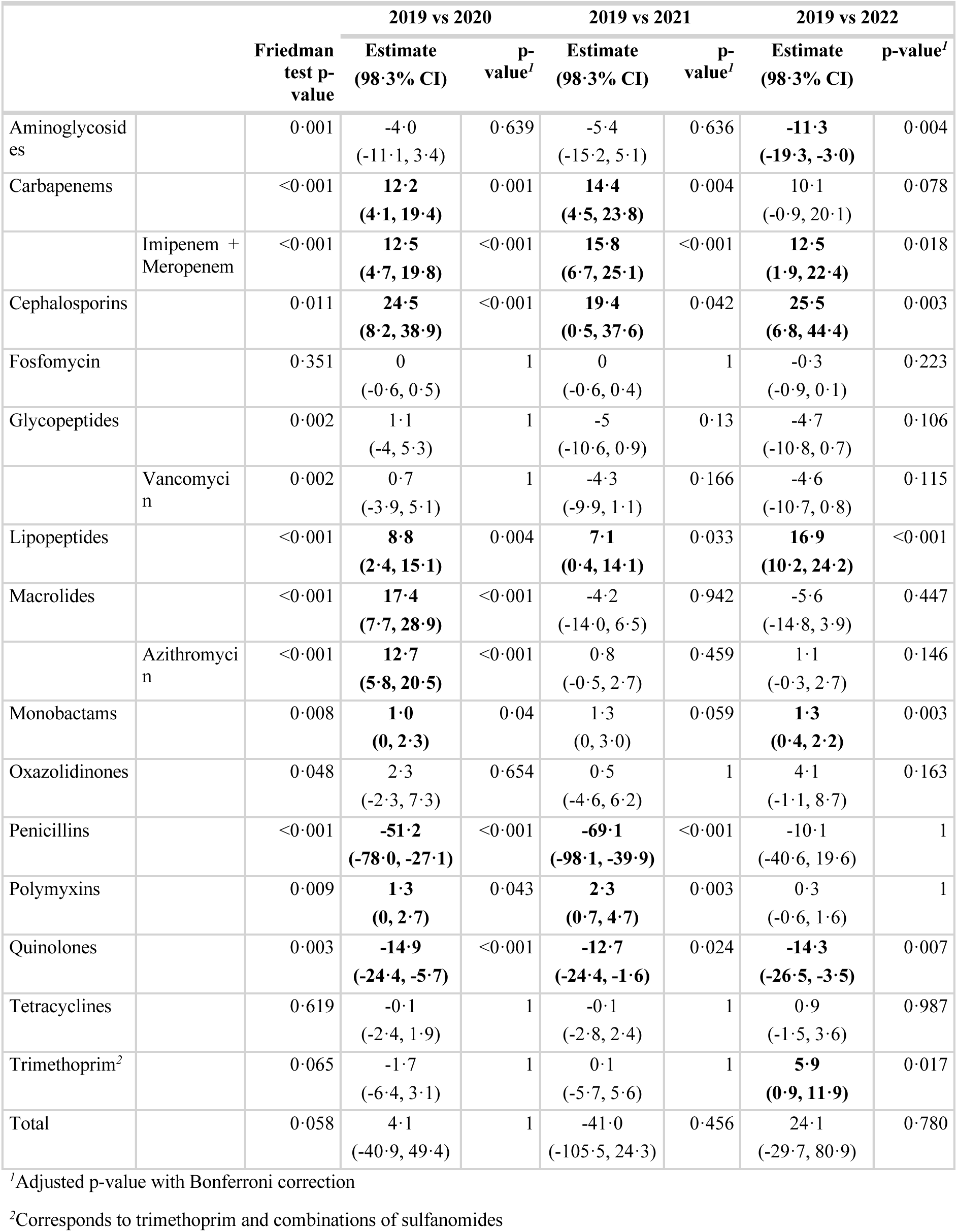
Changes in annual antibiotic consumption in French ICUs, 2019-2022. We report absolute differences in consumption at the ICU level for every antibiotic class, as well as imipenem and meropenem, vancomycin, and azithromycin specifically. Estimates correspond to the median of the differences of antibiotic consumption (in DDD per 1,000 bed-days) between two years of the study period, using 2019 as a reference year. Positive values thereby indicate an increased consumption compared to 2019 and negative values indicate a reduced consumption compared to 2019. For every pairwise comparison, we report the adjusted p-value of the paired Wilcoxon signed-rank test using a Bonferroni correction and the associated 98.3% CI. Finally, we report Friedman tests p-values that are the non- parametric equivalent of one-way repeated measures ANOVA tests to evaluate whether there are changes in antibiotic use over the study period.

### Contrasting impacts of the pandemic on ABR in French hospitals

Among the 692,551 included isolates, *E. coli* was the most represented bacteria in our dataset with 376,685 isolates (Figure 3A). The evolution of annual resistance rates over 2019-2022 varied across antibiotic-bacterium pairs. We found a significantly decreasing trend for MRSA (p<0.001), ESBL-EC (p<0.001), ESBL-KP (p<0.001), and ESBL-ECC (p=0.031), but a significantly increasing trend for CR-PA (p<0.001, Figures 3B-C). No clear variation of resistance proportion at the weekly level was observed (appendix p 8).

**Figure 3.**
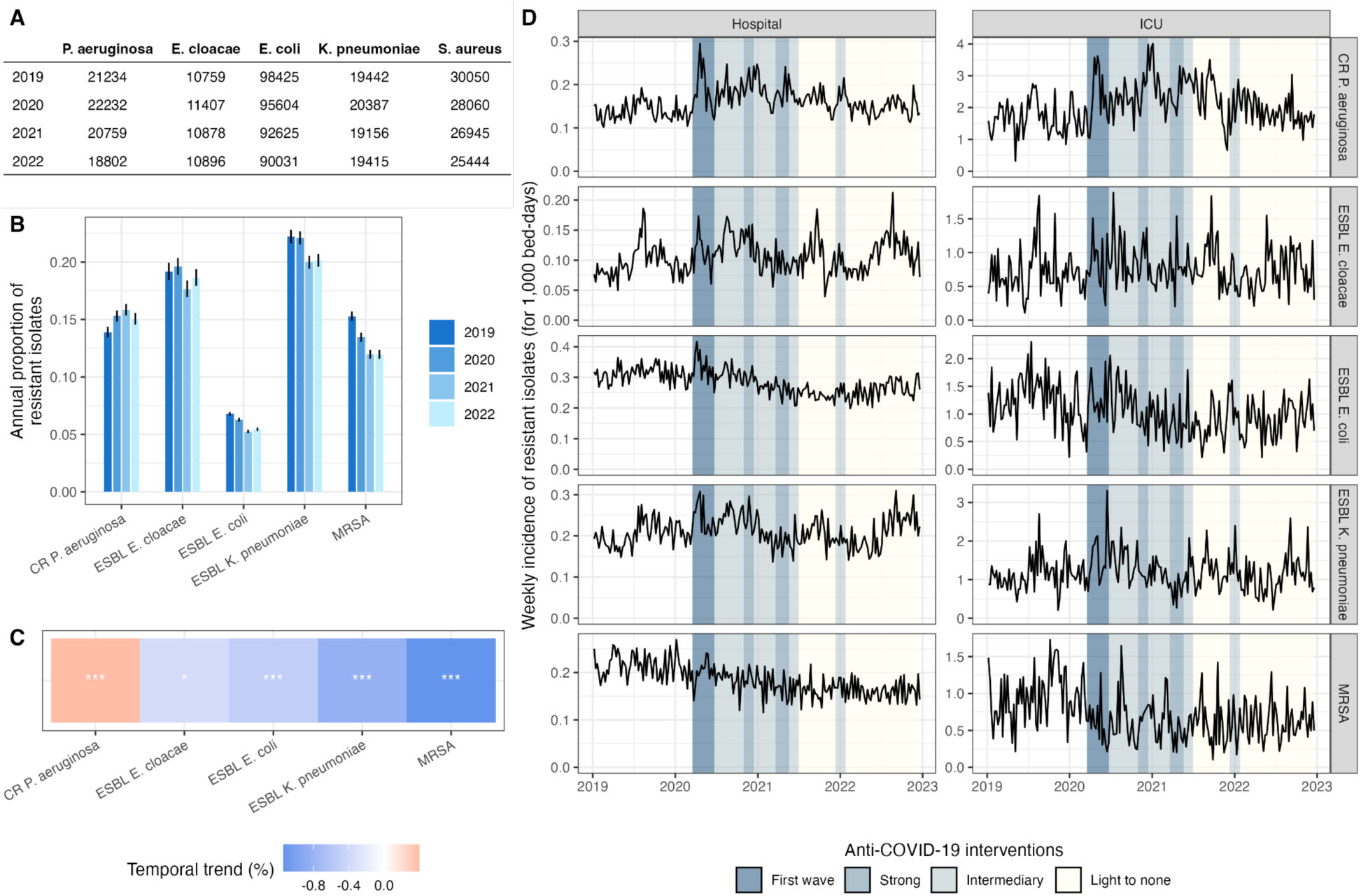
Antibiotic resistance in hospitals in France, 2019-2022. (**A**) Annual number of samples isolated in our hospital cohort (*n*=414 hospitals) between 2019 and 2022 and stratified by bacterial species. (**B**) Annual proportion of resistant bacterial isolates from 2019 to 2022 in our hospital cohort. Intervals indicate the 95% Wilson CIs. (**C**) Temporal trend of resistance proportions between 2019 and 2022. Stars indicate the level of significance of the 𝜒^2^ trend test for proportions. (**D**) Weekly incidence of resistant infections for 1,000 bed-days over the study period in hospitals and intensive care units (ICUs). The strips indicate the level of anti-COVID-19 interventions in the community.^2^ *: p-value≤0.05; **: p-value≤0.01; ***: p-value≤0.001. ESBL E. cloacae: ESBL-producing *Enterobacter cloacae* complex; ESBL E. coli: ESBL-producing *Escherichia coli*; ESBL K. pneumoniae: ESBL-producing *Klebsiella pneumoniae*; CR P. aeruginosa: carbapenem-resistant *Pseudomonas aeruginosa*; MRSA: methicillin-resistant *Staphylococcus aureus*.

In contrast, the analysis of weekly incidence of resistant isolates revealed interesting dynamics (Figure 3D). Comparing regression models that included or not COVID-19-related variables at the country level led to the selection of distinct models depending on the antibiotic-bacterium pair. For ESBL-EC and MRSA, the selected model included the pandemic periods. For CR-PA, the prevalence of intubated COVID-19 patients two weeks prior was selected. Incidence significantly decreased for ESBL-EC (up to 15% decrease, 95% CI: 11-19%) and MRSA (up to 26% decrease, 95% CI: 22-30%) after the first pandemic wave, while the prevalence of intubated COVID-19 patients was significantly associated with higher incidence of CR-PA isolates (p<0.001, Figure 4A).

**Figure 4.**
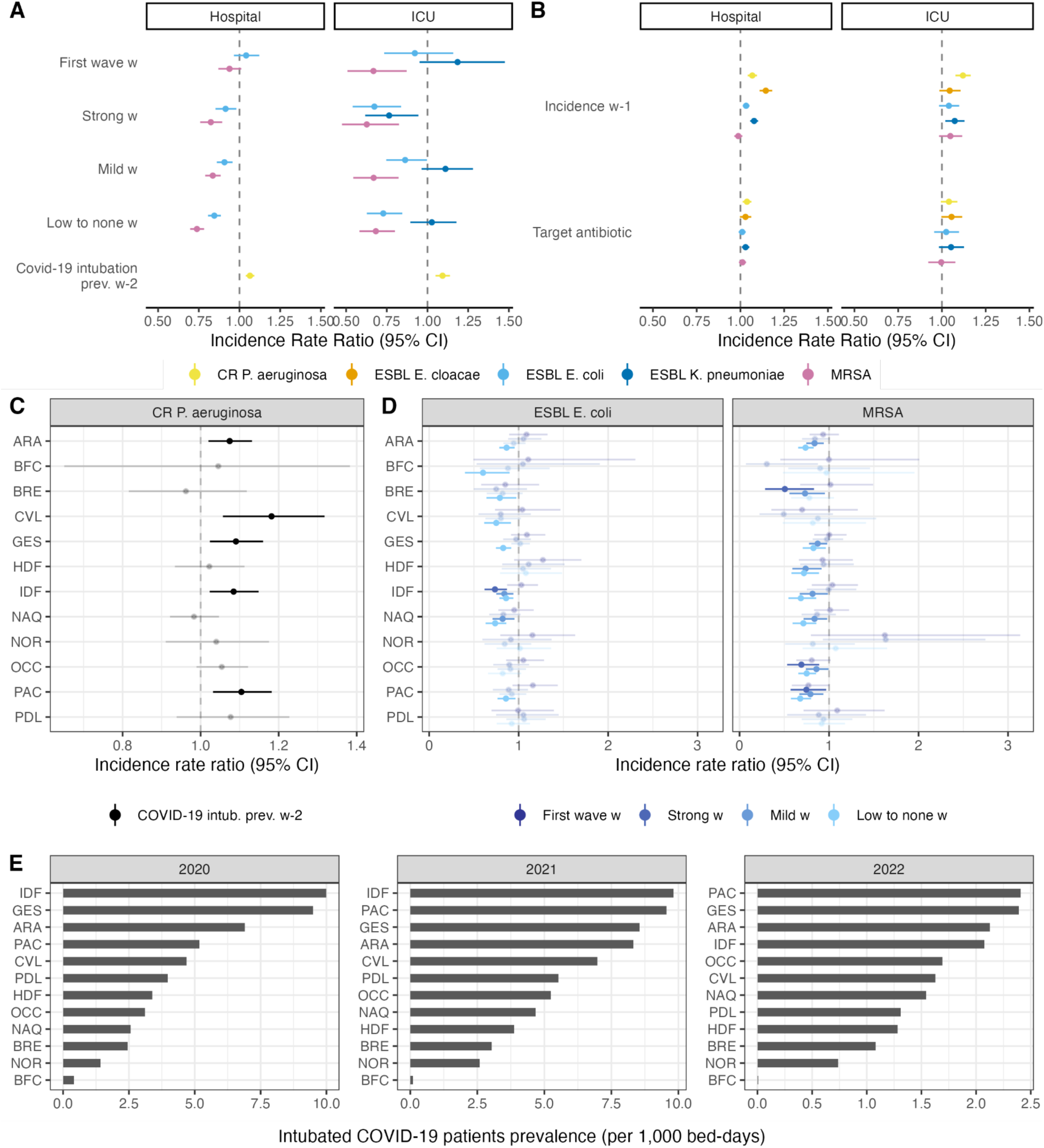
Results from the count regression analysis of resistant infections in French hospitals and intensive care units (ICUs). (**A**) Incidence rate ratios (IRRs) of COVID-19-related variables for the best selected regression models. For ESBL-producing *E. cloacae* in ICUs and hospitals and *K. pneumoniae* in hospitals, the best models did not include COVID-19-related variables, thus they do not appear on the forest plots. For the other cases, the best models included either the COVID-19-related periods at week *w*, or the COVID-19 intubation prevalence at week *w-2*. IRR estimates for the COVID-19-related periods are relative to the pre-pandemic period. (**B**) IRRs of best regression models for the autocorrelation term and antibiotic consumption. Here, “target antibiotic” refers to the antibiotic class targeted by the resistance of the pathogen considered, namely broad-spectrum penicillins for ESBL-producing Enterobacterales, imipenem+meropenem for CR-PA, and narrow spectrum penicillins for MRSA. (**C**) IRRs by administrative region using the best model selected at the national level on carbapenem-resistant *P. aeruginosa* (CR- PA) isolate incidence including the prevalence of COVID-19 intubated patients. (**D**) IRRs by administrative region using the best model at the national level on ESBL-producing *E. coli* (ESBL *E. coli*) and methicillin-resistant *S. aureus* (MRSA) isolate incidence including the pandemic periods at week *w*. Shaded IRRs have a p-value > 0.05. Intervals correspond to the 95% CIs of the point estimates. (**E**) Annual prevalence of intubated COVID-19 patients for 1,000 bed-days by region. Île-de-France (IDF), Grand-Est (GES), Provence-Alpes-Côte d’Azur (PAC), and Auvergne-Rhône-Alpes (ARA) were the most affected regions during the first two years of the pandemic. ESBL E. cloacae: ESBL-producing *Enterobacter cloacae* complex; ESBL E. coli: ESBL-producing *Escherichia coli*; ESBL K. pneumoniae: ESBL-producing *Klebsiella pneumoniae*; CR P. aeruginosa: carbapenem-resistant *Pseudomonas aeruginosa*; MRSA: methicillin-resistant *Staphylococcus aureus*. ARA: Auvergne-Rhône-Alpes; BFC: Bourgogne-Franche-Comté; BRE: Bretagne; CVL: Centre-Val de Loire; GES: Grand-Est; HDF: Hauts-de-France; IDF: Île-de-France ; NAQ: Nouvelle-Aquitaine; NOR: Normandie; OCC: Occitanie; PAC: Provence-Alpes-Côte d’Azur; PDL: Pays de la Loire.

Focusing on ICUs, we observed similar patterns with slight differences (Figure 4A). Notably, MRSA isolates incidence significantly decreased from the first pandemic wave onwards (up to 37% decrease, 95% CI: 18- 53%) and the best model for ESBL-KP included the pandemic periods with a significant decrease in incidence during the periods of strong anti-COVID-19 interventions (24% decrease, 95% CI: 6-38%).

At the regional level, we estimated that COVID-19 intubation prevalence was significantly associated with an increased incidence of CR-PA isolates in IDF (p=0.005), GES (p=0.006), ARA (p=0.008), and PAC (p=0.004, Figure 4C). Interestingly, these regions were amongst the most affected by the pandemic (Figure 4E). These results strengthen the evidence that the pandemic may have led to an increase in CR-PA isolate incidence. For ESBL-EC and MRSA, associations between incidence and pandemic periods are highly heterogeneous across administrative regions with evidence of a significant decrease in incidence after the first pandemic wave in multiple regions (Figure 4D).

## Discussion

Using national surveillance data in French hospitals between 2019 and 2022, we showed changes in antibiotic use over the course of the pandemic, highlighting increasing use of azithromycin, imipenem, and meropenem. We also found that the pandemic had contrasting impacts on ABR, that varied across antibiotic-bacterium pairs and administrative regions, which could be due to differences in the modes of transmission, spatiotemporal dynamics, and drivers of the five studied antibiotic-bacterium pairs. These differences across pairs stress that ABR should not be regarded as a one-solution problem.

We estimated overall reductions in use of penicillins in hospitals over the study period, as observed in 2020 in the community in France,^4^ probably due to the reduction of common infections generally treated with penicillins. They were probably replaced by cephalosporins and carbapenems for which we showed an increased use. We also showed an increased use of macrolides, among which azithromycin, as observed in the community.^3,4^ These changes are certainly due to the management of COVID-19 patients, at least at the start of the pandemic, when COVID-19 patients represented most of the hospitalizations. Indeed, cephalosporins, carbapenems, and azithromycin were the antibiotics most commonly prescribed to COVID-19 patients.^8,12,13^ Importantly, we showed that azithromycin use in hospitals and imipenem and meropenem use in ICUs remained high until 2022. These increases are concomitant with higher carbapenem resistance in *P. aeruginosa* as shown here, and could have led to the increase of azithromycin resistance,^20^ or even other resistances.^20^ Unfortunately, we could not evaluate azithromycin resistance given the very limited number of bacterial isolates tested for this antibiotic. All these dynamics may have changed selective pressures in hospitals and require careful attention. It is important to stress that we did not assess antibiotic use dynamics in the community, which can also impact the selective pressure in hospitals.

To the best of our knowledge, this is the first study evaluating the association between ABR and COVID- 19-related variables in hospitals using weekly level national surveillance data that also include antibiotic use or autocorrelation terms.^10,21^ The results from our regression analysis suggest that CR-PA isolate incidence increased with COVID-19 intubation prevalence, an association that we also quantified at the regional level and that is certainly driven by ICUs (appendix p 26). We further confirmed this association in a sensitivity analysis where likelihood ratio tests always selected the model with COVID-19 intubation at *w* − 2 rather than COVID-19 intubation in the following weeks (*w* + 1, *w* + 2, and *w* + 3, appendix pp 26-27). These results are in agreement with previous studies.^15,16,22^ A possible mechanism explaining this association pertains to the increase of known risk factors of CR-PA infections, notably more frequent intubation,^23^ longer hospitalizations, and higher antibiotic use in COVID-19 patients.^23,24^ Interestingly, when we did not account for imipenem and meropenem use in the hospital regression model, the AICs of the models including the pandemic periods at week *w* − 1 or the prevalence of intubated COVID-19 patients at week *w* − 2 were equivalent. Still, we estimated an increased incidence compared to the pre- pandemic period in the model with the pandemic periods at week *w* − 1(appendix pp 22-25).

In contrast, our analysis shows that MRSA and ESBL-EC isolates incidence significantly decreased during the pandemic. Previous reports have highlighted a steady decrease of MRSA incidence since 2003 in France,^25^ as well as a recent decrease of ESBL-EC between 2016 and 2018.^26^ Thus, it is difficult to assess whether the decrease that we observe is due to long-term trends only or whether the pandemic has accelerated the decrease, for instance through higher adherence to hand hygiene,^5^ a known factor of MRSA prevention.^27^ Concerning ESBL-EC, we estimated a decrease in incidence after the first wave, but our regression model does not explain incidence dynamics well during the first wave (appendix p 19). According to a study in French ICUs during the first wave, COVID-19 patients were more frequently co- infected with ESBL-EC than non-COVID-19 patients^28^ and a meta-analysis highlighted a higher incidence of ESBL-EC during the first year of the pandemic.^15^ It is therefore possible that the impacts of the pandemic on ESBL-EC changed first with conditions that favored ESBL-EC infections, later followed by conditions that prevented these infections. Again, our model did not account for the community and we also cannot rule out the contribution of the evolution of ABR in the community on these dynamics. However, there is scarce evidence of changes in ESBL-EC^29^ and MRSA in the French community.

In parallel, our regression analysis suggests that there is transient association between COVID-19 and ESBL-KP and no association between COVID-19 and ESBL-ECC. In our sensitivity analysis, we estimated stronger changes in ESBL-KP isolate incidence at the start of the pandemic, but they did not persist and concerned only ICUs (appendix pp 18-19). This is consistent with the literature that showed an insignificant increase in the incidence of ESBL-producing Enterobacterales.^15,16^

Several limitations of our study should be mentioned. As in any ecological study, we quantified changes using data aggregated at the national or regional level, which hides hospital heterogeneity in terms of size, hospital type, local practices, or degree to which they were affected by the COVID-19 pandemic. This also prevents generalization to every French hospital. Secondly, the analyses were carried out on a subsample of French hospitals, representing about 14% of all French hospitals (3,008 in 2019). While this cohort is relatively representative in terms of healthcare-associated infections prevalence across regions, it is not representative of the regional distribution of hospital activity or hospital number in France. Besides, hospitals report their data on a voluntary basis. Consequently, we cannot rule out that there is a selection bias in our cohort. For instance, the observed regional differences in antibiotic consumption and CR-PA incidence may be due to missing university hospitals in some regions. Thirdly, the source of acquisition, i.e. whether bacteria were acquired in the community or in the hospital, was not available. Analyzed data thereby result from dynamics of both settings. Fourth, we did not stratify incidence by specimen type (e.g. bloodstream, genital tract, respiratory tract, skin, body fluids, feces, and urine). We might expect different temporal patterns over the year, notably for lower respiratory tract specimens. Fifth, hospitals only report their antibiotic consumption annually, preventing us from exploring weekly level associations with ABR incidence. Indeed, antibiotic use may display seasonal patterns as observed in the community.^30^ Finally, the national surveillance system of ABR changed in 2018, making it impossible to have weekly time series of ABR prior to 2019. Due to this short history, we could not include seasonality in the regression model or account for long-term trends, as previously discussed.

In conclusion, hospital antibiotic use and ABR epidemiology strongly varied during the COVID-19 pandemic in France. The biological mechanisms underpinning the changes in ABR epidemiology likely vary across the investigated antibiotic-bacterium pairs and remain to be elucidated. Continued surveillance efforts in hospitals is pivotal and will help build healthcare facilities that are more resilient in pandemic contexts.

## Contributions

ML, LT, LO, and LW designed the statistical analyses. ML, LT, LO, and SK interpreted the results. ML and DRMS analyzed the data. LS, CD, LD, AJ, SM and LG provided support on data interpretation and analysis. ML, LT, and LO wrote the manuscript. All authors had access to the processed data and reviewed the manuscript. All authors take responsibility for the decision to submit for publication.

## Supporting information

STROBE Checklist

## Data Availability

PMSI and SPARES data are not publicly available. R codes to describe antibiotic consumption and quantify the association between COVID-19-related variables and national or regional resistance data for the antibiotic-bacterium pairs of interest are available online (https://github.com/mlayan/abr_covid_in_french_hospitals).

## Acknowledgments

ML received funding from the National Clinical Research Program and the Investissement d’Avenir program, Laboratoire d’Excellence “Integrative Biology of Emerging Infectious Diseases” (ANR-10-LABX- 62- IBEID). ML thanks the Fondation des Treilles for their support. The authors would like to thank Christian Brun-Buisson for his helpful comments on bacterial sample selection, and Fanny Chéreau for the very insightful discussions on national surveillance systems collecting hospitalizations of COVID-19 patients.

## Ethics approval

Because the analysis was performed using anonymized surveillance data, ethical consent was not required according to the French Data Protection Act. The database was accredited by the French National Data Protection Commission (CNIL no. 2211022 v 0), and the fully anonymized data waiver for informed consent of patients was applied.

