## Supplementary Materials for "Impacts of the COVID-19 pandemic on antibiotic use and resistance in hospitals: a retrospective ecological analysis of French national surveillance data over 2019-2022"

### Table of contents

**1. Antibiotic consumption data in SPARES** **2**
**2. Hospital cohort selection** **3**
**3. Extraction of bed-days data from the PMSI** **6**
**4. Descriptive analysis of antibiotic resistance data** **7**
**5. Statistical modeling** **10**
a. Model selection 10
b. Verification of model assumptions 11
c. Unadjusted estimates 13
**d. Best model fits and estimates** **18**
**6. Evolution in antibiotic use** **20**
**7. Sensitivity analysis exploring the impact of antibiotic use on incidence rate ratios estimates** **22**
**8. Sensitivity analyses for CR-PA isolates incidence** **26**
a. Dynamics of CR-PA isolate incidence in hospitals is driven by intensive care units 26
b. Exploration of the causal relationship between COVID-19 related variables and CR-PA isolate incidence in hospitals and intensive care units 26
**References** **28**

### 1. Antibiotic consumption data in SPARES

Hospital pharmacies report their annual consumption of antiinfectives for systemic use (J01 ATC codes), rifampicin, antiprotozoals, and fidaxomicin to SPARES. We considered the following antibiotic classes: aminoglycosides, carbapenems, cephalosporins, fosfomycin, glycopeptides, lipopeptides, macrolides, monobactams, oxazolidinones, penicillins and their combinations, polymyxins, quinolones, tetracyclines, and trimethoprim and combinations of sulfonamides. We also investigated consumption changes in specific molecules: imipenem and meropenem, vancomycin, and azithromycin. We were interested in imipenem and meropenem as they are the two carbapenems used to treat *P. aeruginosa* infections and their use is a known risk factor of carbapenem-resistant *P. aeruginosa* (CR-PA) infections.[1,2] Vancomycin was also shown to be a risk factor of CR-PA infections.[1] Finally, we were interested in azithromycin consumption given that it has been largely used at the start of the pandemic but was later shown to be ineffective against COVID-19.[3] Azithromycin is also known to promote other resistances.[4]

Among all the molecules or combinations of molecules available in the database (*n*=90), we excluded antibiotics whose ATC codes did not correspond to antiinfectives for systemic use (A07AA12 : fidaxomicin, P01AB01 : metronidazole, P01AB03 : ornidazole, J04AB02 : rifampicin, and P01AB02 : tinidazole), those that did not fall into the antibiotic classes of interest (fusidic acid, erythromycin + sulfafurazole, metronidazole, nitrofurantoin, ornidazole, spiramycin + metronidazole, streptomycin, sulfamethizole, tedizolid, and thiamphenicol), and those that were not reported over the four years of the study period and whose indication is close to anecdotal (cefiderocol, delafloxacin, erythromycin + sulfafurazole, imipenem + relebactam, lincomycin, meropenem + vaborbactam, midecamycin, oritavancin, tedizolid, and telithromycin, Supplementary Figure 1). The ATC codes of included molecules are listed in Supplementary Table 1.

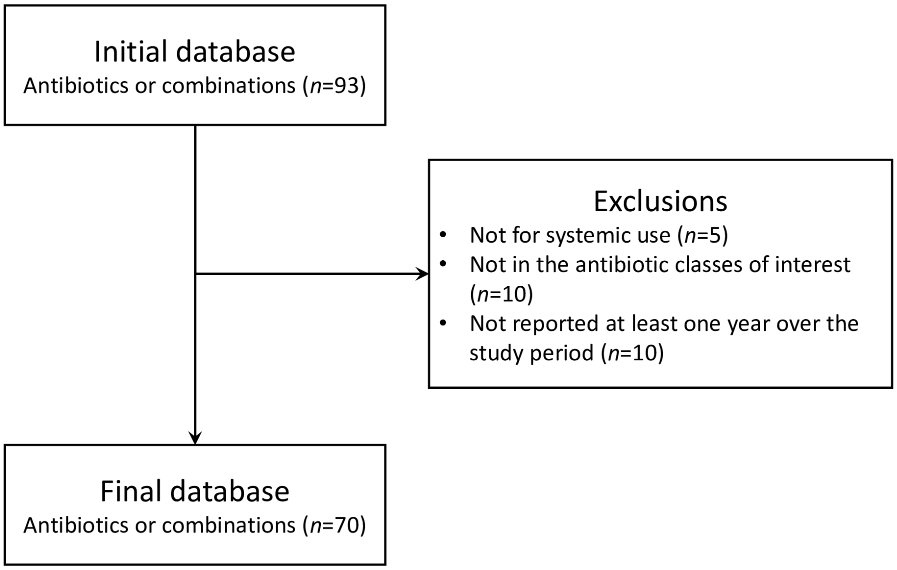


**Supplementary Figure 1. Flow chart of antibiotic molecule selection.**

**Supplementary Table 1. Molecules included in each antibiotic class for antibiotic consumption analyses.** Antibiotic classification is from the WHO/ATC indexing system for antimicrobials (https://www.whocc.no/atc_ddd_index/).

| Antibiotic class | Corresponding group in WHO/ATC indexing system | Molecules |
|---|---|---|
| **Aminoglycosides** | J01GB | Amikacin, Gentamicin, Tobramycin |
| **Carbapenems** | J01DH | Ertapenem, Imipenem, Meropenem |
| **Cephalosporins** | J01DB<br>J01DC<br>J01DD<br>J01DE<br>J01DI | Cefaclor, Cefadroxil, Cefalexin, Cefamandole, Cefazolin, Cefepime, Cefixime, Ceftolozane + Tazobactam, Cefotaxime, Cefotiam, Cefoxitin, Cefpodoxime, Ceftaroline, Ceftazidime, Ceftazidime + Avibactam, Ceftobiprole, Ceftriaxone, Cefuroxime |
| **Fosfomycin** | J01XX | Fosfomycin |
| **Glycopeptides** | J01XA | Dalbavancin, Teicoplanin, Vancomycin |
| **Lipopeptides** | J01XX | Daptomycin |
| **Macrolides** | J01FA<br>J01FF<br>J01FG | Azithromycin, Clarithromycin, Clindamycin, Erythromycin, Josamycin, Pristinamycin, Roxithromycin, Spiramycin |
| **Monobactams** | J01DF | Aztreonam |
| **Oxazolidinones** | J01XX | Linezolid |
| **Penicillins** | J01CA<br>J01CE<br>J01CF<br>J01CR | Amoxicillin + Clavulanic acid, Amoxicilline, Ampicillin, Ampicillin + Sulbactam, Benzathine, Benzylpenicillin, Cloxacillin, Piperacillin, Piperacillin + Tazobactam, Oxacillin, Penicillin V, Pivmecillinam, Temocillin, Ticarcillin, Ticarcillin + Clavulanic acid |
| **Polymyxins** | J01XB | Colistin |
| **Quinolones** | J01MA | Ciprofloxacin, Levofloxacin, Lomefloxacin, Moxifloxacin, Norfloxacin, Ofloxacin |
| **Tetracyclines** | J01AA | Demeclocycline, Doxycycline, Lymecycline, Metacycline, Minocycline, Tigecycline |
| **Trimethoprim and combinations of sulfonamides** | J01EA<br>J01EC<br>J01EE | Cotrimoxazole, Sulfadiazine, Trimethoprim |

### 2. Hospital cohort selection

Hospitals report every year their antibiotic resistance (ABR) and antibiotic consumption data to the national surveillance system SPARES on a voluntary basis. Every hospital in France is identified with two FINESS codes: one code designates the geographical/physical entity, and the other designates the legal entity. Every physical entity is associated with a unique legal FINESS code, but a legal FINESS code generally encompasses multiple physical entities.

In SPARES, hospitals report their data using their physical or their legal FINESS code. We did not consider hospitals reporting their data under their legal FINESS code as we expect practices to differ between physical entities. However, for university hospitals, we included hospitals reporting their data using their physical or legal FINESS code (Supplementary Figure 2A) when they met all the other inclusion criteria. This choice was motivated by the key role that university hospitals play in ABR epidemiology in France. Indeed, they generally handle the sickest patients and thereby are central in ABR associated with healthcare. Besides, they were at the frontline during the COVID-19 pandemic which makes them a good setting to evaluate the impacts of the pandemic on ABR.

From the reporting hospitals, we constituted a cohort of hospitals and a nested cohort of ICUs to investigate the impacts of the COVID-19 pandemic on ABR and antibiotic consumption. The distribution of hospital type in our cohort varies a lot across regions (Supplementary Figure 2B).

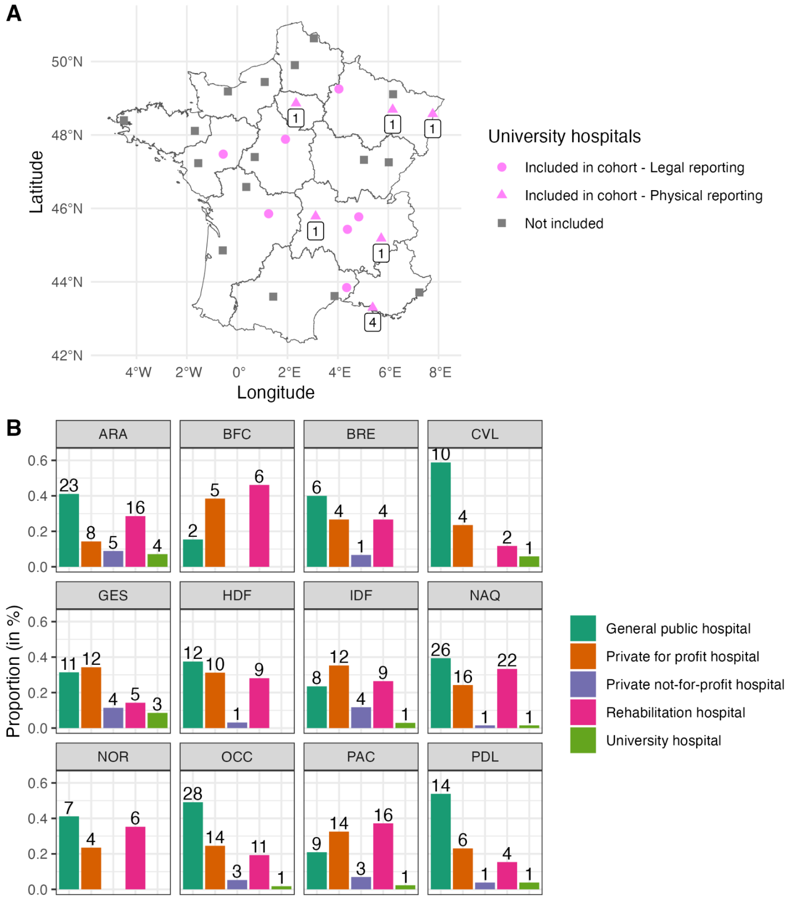


**Supplementary Figure 2. Geographical and hospital type description of our hospital cohort.** (**A**) Physical and legal entities corresponding to university hospitals and included in our cohort. In total, there are 29 legal university hospitals in France. We included only 13 of them (in pink), 7 of which reported data using their legal FINESS code (pink circles). For the remaining included hospitals, only some physical entities reported their data (numbers indicated below the pink triangles). (**B**) Distribution of hospital types by region. Bars indicate the proportion of each hospital type within each region, and numbers correspond to the number of physical entities, except for university hospitals for which we report the number of legal entities. ARA: Auvergne-Rhône-Alpes; BFC: Bourgogne-Franche-Comté; BRE: Bretagne; CVL: Centre-Val de Loire; GES: Grand-Est; HDF: Hauts-de-France; IDF: Île-de-France ; NAQ: Nouvelle-Aquitaine; NOR: Normandie; OCC: Occitanie; PAC: Provence-Alpes-Côte d'Azur; PDL: Pays de la Loire.

To assess the generalizability of our results, we explored the representativity of our cohort. Multiple dimensions of representativity can be explored, notably regional distribution and hospital type distribution, two factors that are expected to shape (*i*) the burden of ABR, (*ii*) healthcare workers (HCW) practices related to infection prevention and control measures and antibiotic prescribing, and (*iii*) the burden and adaptation to the COVID-19 pandemic.

National statistics on hospital types are not available; thereby, we could not assess the representativity of our cohort in terms of hospital type. Other national statistics on hospitals by region are available however, notably the number of acute-care hospitals, the hospital activity in bed-days, and the prevalence of hospital-acquired infections (HAI, Supplementary Figure 3). When comparing the number of legal hospitals and hospital activity in terms of bed-days in 2020 to the national reference, we showed that some regions, notably Île-de-France (IDF) were underrepresented in our sample. The representativity in terms of prevalence of infections could be assessed by comparing the prevalence of bacterial isolates in our hospital cohort and the HAI prevalence from the national point prevalence survey. We could only compare the distributions visually, with a large under estimation in our cohort in Provence-Alpes-Côte d'Azur (PAC).

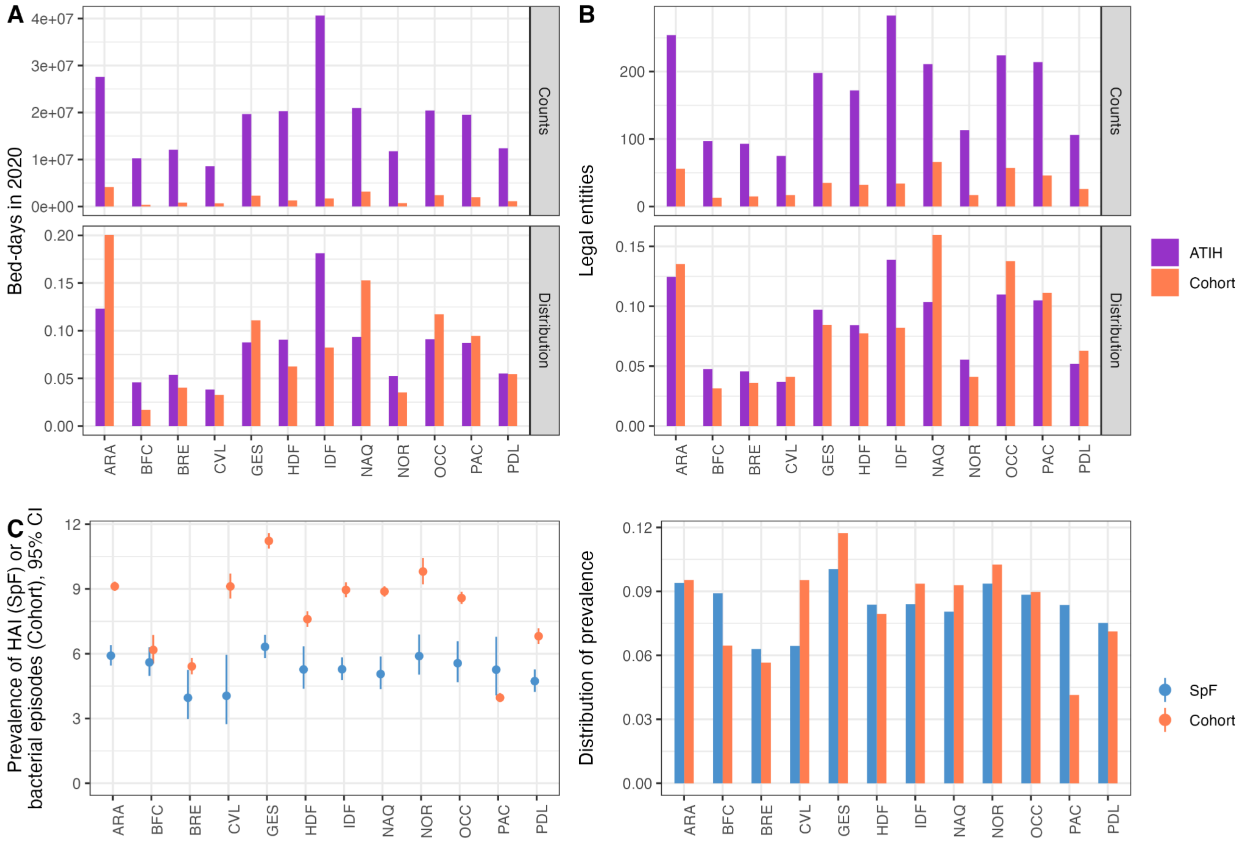


**Supplementary Figure 3. Hospital cohort representativity.** (**A**) Comparison of the distribution of the number of legal entities by region. We compared the regional distribution in our cohort to the distribution reported by the French technical agency on hospitalization information in 2020 (ATIH) using a $\chi^2$ test. The p-value of this test is <0.001 which suggests that our hospital cohort is not representative, notably due to the underrepresentation of hospitals in Île-de-France (IDF). (**B**) Comparison of the distribution of the number of bed-days in 2020 by region. We compared the regional distribution in our cohort to the distribution reported by the French technical agency on hospitalization information in 2020 (ATIH) using a $\chi^2$ test. The p-value of this test is lower than 1e-3 which suggests that our hospital cohort is not representative of hospital activity across regions. It is presumably due to the heterogeneous inclusion of university hospitals across regions (Supplementary Figure 2A). For example, only a minor hospital of the Parisian University Hospital (Assistance publique - Hôpitaux de Paris, AP-HP) is included in our cohort while AP-HP is the largest hospital group in France in terms of activity. (**C**) Comparison of the distribution of the prevalence of healthcare-associated infections (HAIs) at the regional level. We compared the point prevalence of all bacterial and viral HAIs (except SARS-CoV-2 HAIs) estimated by the French national agency of Public health (Santé publique France, SpF) to the prevalence of patients with bacterial infections based on our hospital cohort. The point prevalence estimated by SpF is based on the number of hospitalized patients with a bacterial or viral HAI in participating hospitals over one day between the 15th of May and the 30th of June 2022. In our cohort, we divided the number of patients with a bacterial infection detected between the 15th of May and the 30th of June 2022 and divided it by the number of patients hospitalized in the corresponding hospitals over the same period that we extracted from the PMSI. We have a relatively good agreement between our cohort and SpF, but with an overestimation in Centre-Val de Loire (CVL) and underestimation in Provence-Alpes-Côte d'Azur (PAC). ARA: Auvergne-Rhône-Alpes; BFC: Bourgogne-Franche-Comté; BRE: Bretagne; CVL: Centre-Val de Loire; GES: Grand-Est; HDF: Hauts-de-France; IDF: Île-de-France ; NAQ: Nouvelle-Aquitaine; NOR: Normandie; OCC: Occitanie; PAC: Provence-Alpes-Côte d'Azur; PDL: Pays de la Loire.

#### 3. Extraction of bed-days data from the PMSI

We extracted the weekly number of bed-days and the weekly number of intubated COVID-19 patient bed-days for hospitals of our cohort from the National Hospital Discharge Database (PMSI). Of note, the PMSI has been designed to describe hospital activity in a standard manner and not to conduct epidemiological studies so its use in epidemiological studies has inherent limitations.[5]

For each hospital in our cohort, we used its physical FINESS code to retrieve all patient stays in the PMSI. For each university hospital that reported its data using its legal FINESS code, we listed the associated physical FINESS codes corresponding to acute-care facilities that are publicly available in the national register of FINESS codes (https://www.data.gouv.fr/fr/). We downsampled patient stays to the ones corresponding to the patient population followed-up by SPARES (inpatients) and our selection criteria (individuals above 15 years old, Supplementary Figure 4A).

To extract intubated COVID-19 patient bed-days (Supplementary Figure 4B), we selected from the line list of patient stays above-mentioned COVID-19 patients that were intubated. We identified COVID-19 patients using the U07 and U10 codes of the International Classification of Diseases 10th revision. Tracheal intubation was identified using the GELD004, GELD002, GLLD004 and GLLD008 codes of the French medical classification for clinical procedures (CCAM). The exact date of the diagnosis is not available in the PMSI, thereby we assumed that patients had COVID-19 during their whole stay. For the procedures, the date of the procedure is available but not its duration. We assumed that patients were intubated from the start of the procedure until the end of their stay. This assumption likely overestimates the weekly prevalence of intubated COVID-19 patients as it is very probable that patients are not desintubated a few days prior to the end of their stay/transfer to another medical ward.

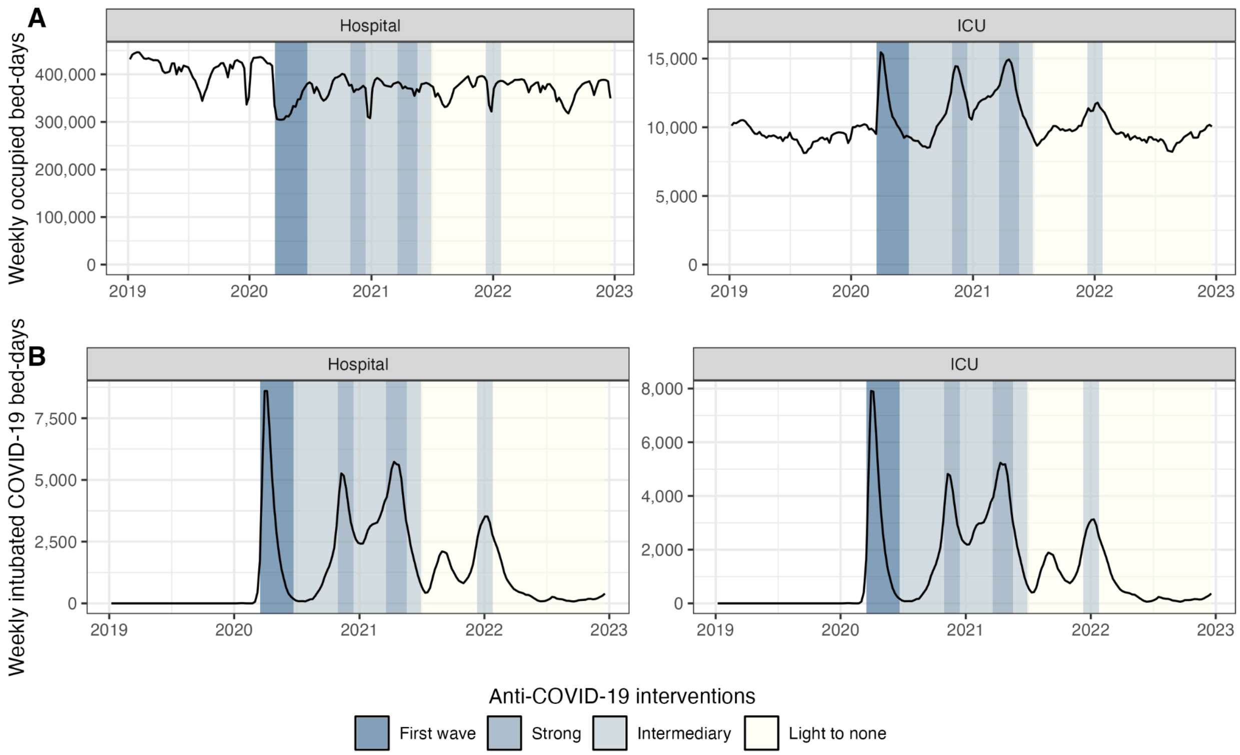


**Supplementary Figure 4. Dynamics of occupied bed-days in hospitals and intensive care units (ICUs) over the study period.** (**A**) Weekly number of occupied bed-days in hospitals and ICUs of our national cohorts. We can notice a seasonal pattern with decreased bed-days during summer and Christmas holidays in 2019. This pattern was largely affected during the pandemic. (**B**) Weekly number of COVID-19 bed-days in hospitals and ICUs of our national cohorts. The strips indicate the level of anti-COVID-19 interventions in the community that we adapted from Paireau and colleagues.[6]

### 4. Descriptive analysis of antibiotic resistance data

We extracted the incident number of episodes caused by five bacterial species (*P. aeruginosa*, *E. cloacae* complex, *E. coli*, *K. pneumoniae*, and *S. aureus*) and classified them as resistant or not. We considered bacteria to be resistant when they had an R phenotype or produced extended-spectrum β-lactamase (BLSE). We obtained the weekly proportion of resistant episodes by dividing the incident number of resistant episodes by the incident number of all episodes. Resistance proportion displayed little short-term dynamics over the study period, except for CR-PA (Supplementary Figure 5).

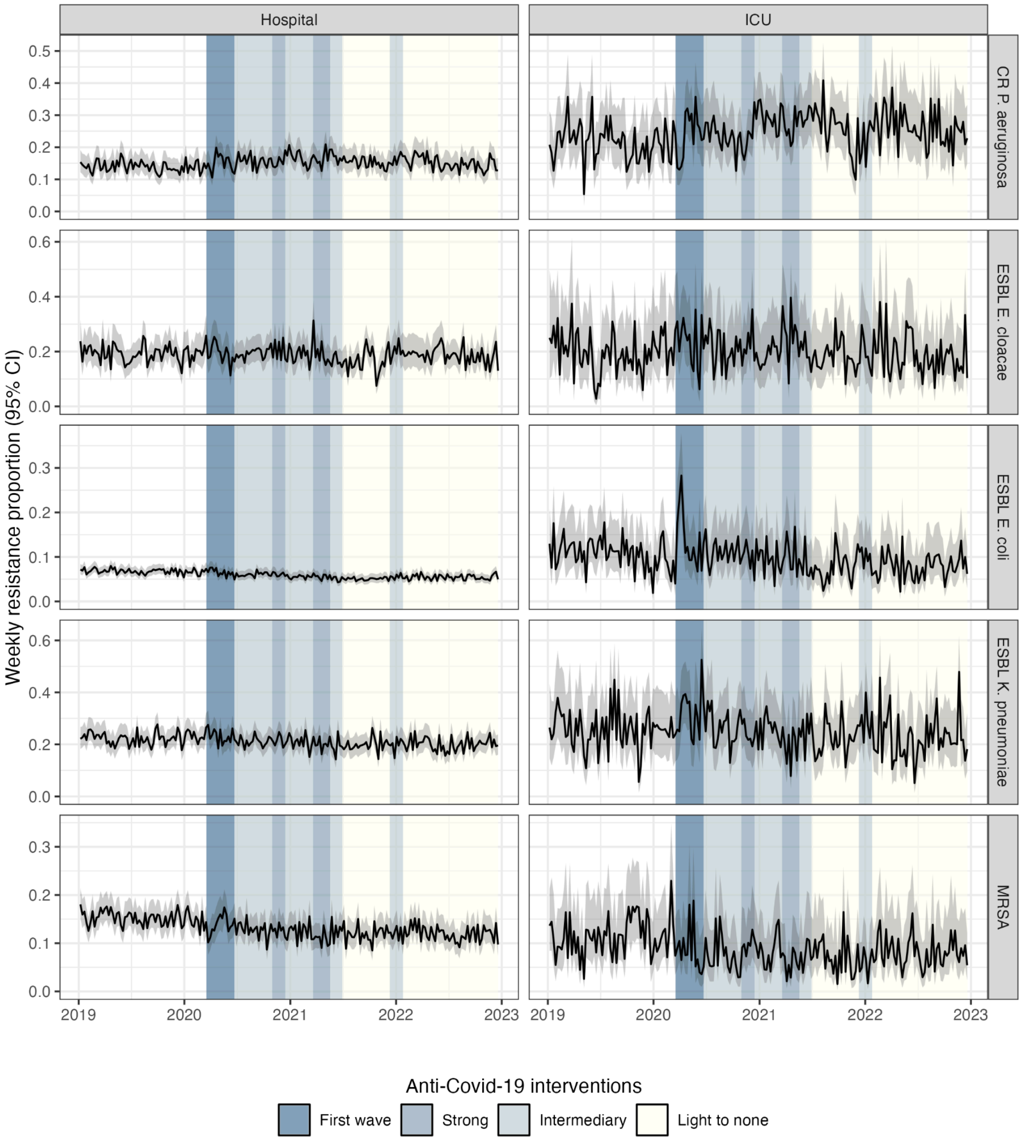


**Supplementary Figure 5. Weekly resistance proportions of bacterial samples isolated in our hospital cohort, 2019-2022.** We calculated the Wilson binomial proportion confidence interval (in grey).

The incidence of resistant isolates displayed more pronounced variations during the study period (Figure 3D). Except for ESBL-producing *E. cloacae* complex (ESBL-ECC) in ICUs, the incidence of the five drug-bacterium pairs significantly changed during the study period (Supplementary Figure 6A). We observed a clear decrease after the start of the pandemic for MRSA, while an increase for CR-PA during the first wave followed by a decrease in the later stages of the pandemic (low to no restrictions). In addition, there is a significant correlation at the hospital and ICU levels between CR-PA incidence and the prevalence of intubated COVID-19 patients (Supplementary Figure 6B).

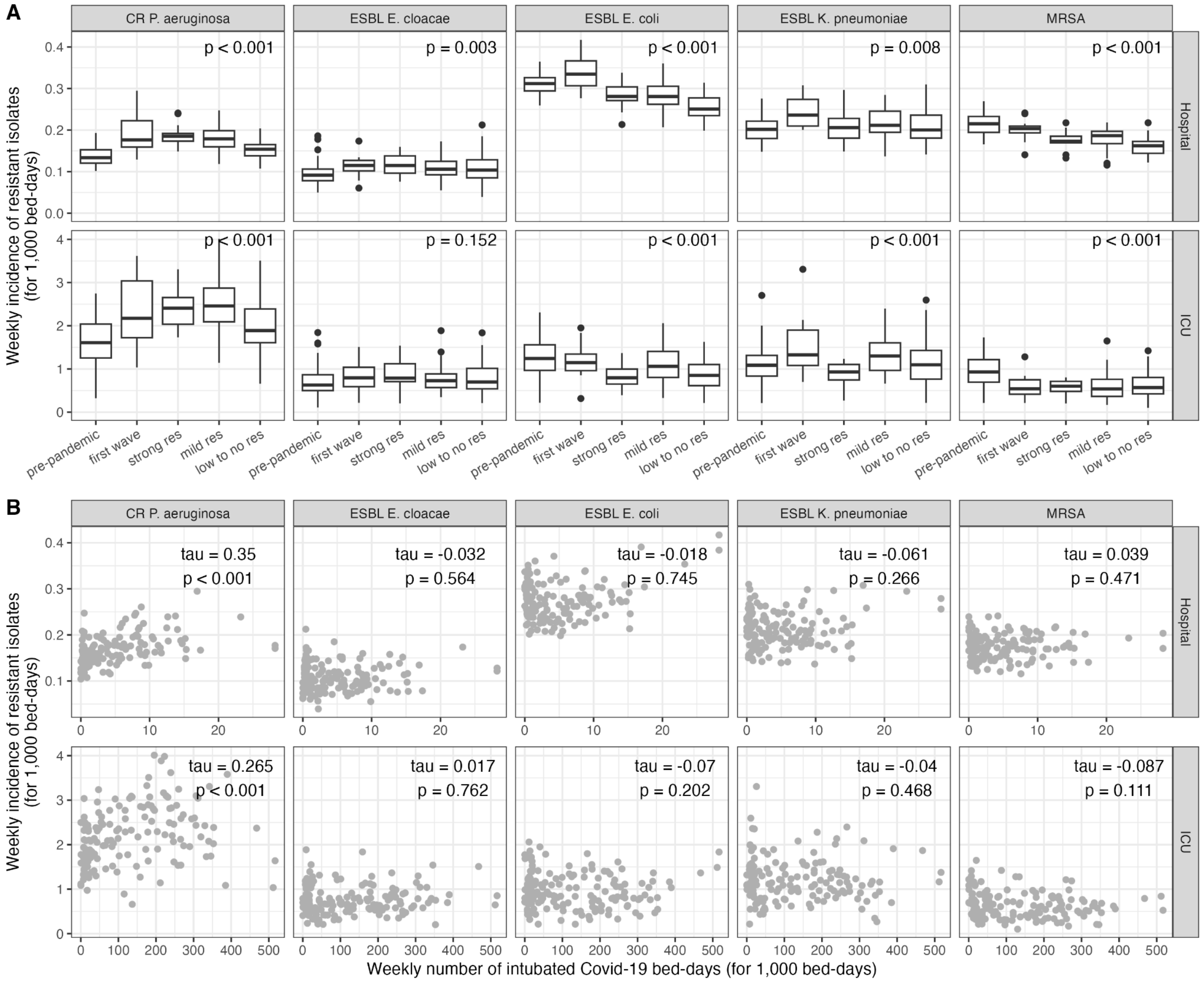


**Supplementary Figure 6. Correlation between the weekly incidence of resistant infections and COVID-19-related variables in hospitals and ICUs at the national level.** (**A**) Variations of the weekly incidence of resistant infections according to the level of anti-COVID-19 restrictions in hospitals and ICUs. The p-value of the Kruskal-Wallis test is indicated on each panel. (**B**) Correlation between the weekly incidence of resistant infections and the weekly prevalence of COVID-19 patients in hospitals and ICUs for the resistant pathogens of interest. Kendall's tau correlation coefficient and the p-value of the correlation test are indicated on each panel.

### 5. Statistical modeling

#### a. Model selection

We tested a baseline model of resistant isolate incidence that did not account for the COVID-19 pandemic and compared it to six models including either the pandemic periods or the prevalence of intubated COVID-19 patients for weeks *w*, *w-1* or *w-2* (Supplementary Table 2).

**Supplementary Table 2. COVID-19-related variables included in the count regression models of infections at the national level.**

| Model | COVID-19-related variables | Additional term in model equation |
|---|---|---|
| **Baseline** | None | - |
| **1** | A categorical variable indicating the COVID-19-related period of week *w*: pre-pandemic (reference), first wave, strong restrictions, intermediate restrictions, low to no restrictions | $\beta \cdot Period_{w}$ |
| **2** | A categorical variable indicating the COVID-19-related period of week *w-1*: pre-pandemic (reference), first wave, strong restrictions, intermediate restrictions, low to no restrictions | $\beta \cdot Period_{w-1}$ |
| **3** | A categorical variable indicating the COVID-19-related period of week *w-2*: pre-pandemic (reference), first wave, strong restrictions, intermediate restrictions, low to no restrictions | $\beta \cdot Period_{w-2}$ |
| **4** | Prevalence of intubated COVID-19 cases (for 1,000 bed-days) of week *w* | $\beta \cdot Prev_{intub-covid,w}$ |
| **5** | Prevalence of intubated COVID-19 cases (for 1,000 bed-days) of week *w-1* | $\beta \cdot Prev_{intub-covid,w-1}$ |
| **6** | Prevalence of intubated COVID-19 cases (for 1,000 bed-days) of week *w-2* | $\beta \cdot Prev_{intub-covid,w-2}$ |

We selected the best model based on the Akaike Information Criterion (AIC, Supplementary Table 3). If no model with COVID-19 variables had an AIC difference of at least 5 compared to the baseline model (without COVID-19 variables), we considered the baseline model as the best model. If multiple COVID-19 models had similar AIC values, we kept the model that would facilitate results presentation. For instance, we used the "Pandemic periods *w*" model for ESBL-producing *K. pneumoniae* (ESBL-KP) in ICUs although it had a higher AIC than the "Pandemic periods *w-1*" model so that we could present the estimates for ESBL-KP, ESBL-producing *E. coli*, and MRSA all at once.

**Supplementary Table 3. Results of the model comparison and AIC values of the regression models tested on the national infection data.** The selected model (AIC in bold) is the one minimizing the AIC.

| | **CR *P. aeruginosa*** | **ESBL-producing *K. pneumoniae*** | **ESBL-producing *E. cloacae* complex** | **ESBL-producing *E. coli*** | **MRSA** |
|---|---|---|---|---|---|
| **Hospital** | | | | | |
| No COVID-19 variable | 1496 | **1601** | **1486** | 1630 | 1570 |
| Pandemic periods *w* | 1485 | 1607 | 1492 | **1585** | **1491** |
| Pandemic periods *w-1* | 1482 | 1607 | 1492 | 1591 | 1491 |
| Pandemic periods *w-2* | 1482 | 1608 | 1492 | 1591 | 1492 |
| COVID-19 intubation prevalence *w* | 1484 | 1603 | 1487 | 1632 | 1571 |
| COVID-19 intubation prevalence *w-1* | 1479 | 1603 | 1487 | 1631 | 1572 |
| COVID-19 intubation prevalence *w-2* | **1476** | 1603 | 1487 | 1630 | 1572 |
| **ICU** | | | | | |
| No COVID-19 variable | 1291 | 1187 | **1051** | 1162 | 1043 |
| Pandemic periods *w* | 1293 | **1181** | 1055 | **1147** | **1027** |
| Pandemic periods *w-1* | 1290 | 1179 | 1056 | 1148 | 1027 |
| Pandemic periods *w-2* | 1287 | 1182 | 1054 | 1149 | 1029 |
| COVID-19 intubation prevalence *w* | 1289 | 1188 | 1053 | 1156 | 1041 |
| COVID-19 intubation prevalence *w-1* | 1283 | 1188 | 1052 | 1156 | 1040 |
| COVID-19 intubation prevalence *w-2* | **1275** | 1189 | 1052 | 1154 | 1040 |

#### b. Verification of model assumptions

**Supplementary Table 4. Dispersion parameter of the negative binomial estimated in regression model analyses.** We report the estimates of the overdispersion parameter with its 95% CI.

| | **Best model** | **Overdispersion (95% CI)** |
|---|---|---|
| **Hospital** | | |
| CR *P. aeruginosa* | COVID-19 intubation prevalence *w-2* | 214 (29, 400) |
| ESBL-producing *E. cloacae* complex | No COVID-19 variable | 39 (24, 54) |
| ESBL-producing *E. coli* | Pandemic periods *w* | 708 (-322, 1737) |
| ESBL-producing *K. pneumoniae* | No COVID-19 variable | 106 (58, 154) |
| MRSA | Pandemic periods *w* | 492 (-254, 1238) |
| **ICU** | | |
| CR *P. aeruginosa* | COVID-19 intubation prevalence *w-2* | 56 (15, 97) |
| ESBL-producing *E. cloacae* complex | No COVID-19 variable | 23 (6, 40) |
| ESBL-producing *E. coli* | Pandemic periods *w* | 25 (8, 41) |
| ESBL-producing *K. pneumoniae* | Pandemic periods *w* | 24 (10, 37) |
| MRSA | Pandemic periods *w* | 31 (-2, 65) |

**Supplementary Table 5. Multicollinearity between variables of the restriction levels of anti-COVID-19 interventions and COVID-19 prevalence.** Here, we report the variance inflation factor (VIF) for all covariables when we include both the prevalence of intubated COVID-19 patients and pandemic periods. Generally, we consider that there is little to no correlation between covariables for VIF values ≤ 5. Values greater than 5 (in bold) are indicative of multicollinearity. We did not explore models with both COVID-19 variables as they often led to VIF values close to 5 or greater than 5.

| | CR *P. aeruginosa* | ESBL-producing *K. pneumoniae* | ESBL-producing *E. cloacae* complex | ESBL-producing *E. coli* | MRSA |
|---|---|---|---|---|---|
| **Hospital** | | | | | |
| Incidence *w-1* | 1·6 | 1·1 | 1·1 | 1·8 | 2·0 |
| COVID-19 intubation prev. *w* | 2·5 | 2·5 | 2·6 | 2·5 | 2·8 |
| Pandemic periods | 4·6 | 4·4 | 4·7 | **7·6** | 5·0 |
| Imipenem + Meropenem | 2·3 | - | - | - | - |
| 3rd generation Cephalosporins | - | 2·3 | 2·3 | 2·2 | - |
| Penicillins | - | - | - | - | 1·3 |
| **ICU** | | | | | |
| Incidence *w-1* | 1·2 | 1·1 | 1·0 | 1·2 | 1·3 |
| COVID-19 intubation prev. *w* | 2·7 | 2·8 | 3·0 | 3·0 | 3·1 |
| Pandemic periods | **6·1** | **5·4** | **5·6** | **6·0** | 4·2 |
| Imipenem + Meropenem | 3·2 | - | - | - | - |
| 3rd generation Cephalosporins | - | 2·2 | 2·2 | 2·1 | - |
| Penicillins | - | - | - | - | 1·9 |

**Supplementary Table 6. Residuals autocorrelation of the best count regression models selected in the national analysis.** We performed Box-Ljung tests on 52 lags to evaluate whether residuals of the negative binomial regression model were autocorrelated. A p-value greater than 0·05 indicates that residuals are not autocorrelated. At the hospital level, the best models fail to account for the autocorrelation over 52 weeks, except for MRSA. At the ICU level, there is no autocorrelation in the residuals over 52 weeks, except for ESBL-KP and ESBL-EC.

| | Chi2 | df | p.value |
|---|---|---|---|
| **Hospital** | | | |
| CR *P. aeruginosa* | 74·26 | 51 | 0·018 |
| ESBL-producing *E. cloacae* complex | 98·85 | 51 | <0·001 |
| ESBL-producing *E. coli* | 83·52 | 51 | 0·003 |
| ESBL-producing *K. pneumoniae* | 72·04 | 51 | 0·028 |
| MRSA | 51·51 | 51 | 0·454 |
| **ICU** | | | |
| CR *P. aeruginosa* | 56·51 | 51 | 0·277 |
| ESBL-producing *E. cloacae* complex | 45·20 | 51 | 0·702 |
| ESBL-producing *E. coli* | 69·73 | 51 | 0·042 |
| ESBL-producing *K. pneumoniae* | 94·52 | 51 | <0·001 |
| MRSA | 59·66 | 51 | 0·190 |

#### c. Unadjusted estimates

**Supplementary Table 7. Unadjusted estimates of incidence rate ratios (IRRs) associated to COVID-19-related variables for CR-PA in French hospitals and intensive care units (ICUs).** We depicted in bold the IRRs whose p-value is lower than 0·05.

| **CR *P. aeruginosa*** | | **IRR** | **p-value** |
|---|---|---|---|
| **Hospital** | | | |
| **Pandemic periods *w*** | | | **<0·001[1]** |
| | First wave | 1·39 (1·27, 1·52) | **<0·001** |
| | Strong res | 1·38 (1·26, 1·50) | **<0·001** |
| | Mild res | 1·33 (1·25, 1·41) | **<0·001** |
| | Low to no res | 1·11 (1·05, 1·17) | **<0·001** |
| **Pandemic periods *w-1*** | | | **<0·001[1]** |
| | First wave | 1·38 (1·26, 1·51) | **<0·001** |
| | Strong res | 1·44 (1·32, 1·57) | **<0·001** |
| | Mild res | 1·32 (1·24, 1·40) | **<0·001** |
| | Low to no res | 1·11 (1·05, 1·17) | **<0·001** |
| **Pandemic periods *w-2*** | | | **<0·001[1]** |
| | First wave | 1·37 (1·25, 1·50) | **<0·001** |
| | Strong res | 1·45 (1·33, 1·58) | **<0·001** |
| | Mild res | 1·31 (1·24, 1·39) | **<0·001** |
| | Low to no res | 1·10 (1·04, 1·16) | **<0·001** |
| **COVID-19 intubation prevalence** | Week *w* | 1·11 (1·08, 1·13) | **<0·001** |
| | Week *w-1* | 1·12 (1·09, 1·14) | **<0·001** |
| | Week *w-2* | 1·12 (1·10, 1·15) | **<0·001** |
| **ICU** | | | |
| **Pandemic periods *w*** | | | **<0·001[1]** |
| | First wave | 1·38 (1·17, 1·63) | **<0·001** |
| | Strong res | 1·43 (1·22, 1·68) | **<0·001** |
| | Mild res | 1·48 (1·32, 1·66) | **<0·001** |
| | Low to no res | 1·20 (1·08, 1·34) | **<0·001** |
| **Pandemic periods *w-1*** | | | **<0·001[1]** |
| | First wave | 1·36 (1·15, 1·60) | **<0·001** |
| | Strong res | 1·53 (1·31, 1·79) | **<0·001** |
| | Mild res | 1·47 (1·31, 1·64) | **<0·001** |
| | Low to no res | 1·18 (1·07, 1·32) | **0·002** |
| **Pandemic periods *w-2*** | | | **<0·001[1]** |
| | First wave | 1·43 (1·22, 1·68) | **<0·001** |
| | Strong res | 1·64 (1·41, 1·91) | **<0·001** |
| | Mild res | 1·45 (1·30, 1·62) | **<0·001** |
| | Low to no res | 1·19 (1·07, 1·31) | **0·001** |
| **COVID-19 intubation prevalence** | Week *w* | 1·12 (1·08, 1·17) | **<0·001** |
| | Week *w-1* | 1·15 (1·10, 1·19) | **<0·001** |
| | Week *w-2* | 1·17 (1·13, 1·21) | **<0·001** |

[1]Global p-value

**Supplementary Table 8. Unadjusted estimates of incidence rate ratios (IRRs) associated to COVID-19-related variables for ESBL-producing *E. cloacae* complex in French hospitals and intensive care units (ICUs).** We depicted in bold the IRRs whose p-value is lower than 0·05.

| **ESBL-producing *E. cloacae* complex** | | **IRR** | **p-value** |
|---|---|---|---|
| **Hospital** | | | |
| **Pandemic periods *w*** | | | **0·012**[1] |
| | First wave | 1·18 (1·01, 1·38) | **0·034** |
| | Strong res | 1·21 (1·05, 1·41) | **0·011** |
| | Mild res | 1·16 (1·05, 1·28) | **0·004** |
| | Low to no res | 1·12 (1·02, 1·22) | **0·013** |
| **Pandemic periods *w-1*** | | | **0·021**[1] |
| | First wave | 1·16 (1·00, 1·36) | 0·051 |
| | Strong res | 1·16 (0·99, 1·34) | 0·060 |
| | Mild res | 1·17 (1·06, 1·29) | **0·002** |
| | Low to no res | 1·12 (1·02, 1·22) | **0·014** |
| **Pandemic periods *w-2*** | | | **0·013**[1] |
| | First wave | 1·19 (1·03, 1·39) | **0·022** |
| | Strong res | 1·12 (0·96, 1·30) | 0·150 |
| | Mild res | 1·17 (1·06, 1·30) | **0·001** |
| | Low to no res | 1·13 (1·03, 1·23) | **0·009** |
| **COVID-19 intubation prevalence** | Week *w* | 1·03 (0·99, 1·06) | 0·168 |
| | Week *w-1* | 1·03 (0·99, 1·07) | 0·139 |
| | Week *w-2* | 1·02 (0·99, 1·06) | 0·259 |
| **ICU** | | | |
| **Pandemic periods *w*** | | | 0·208[1] |
| | First wave | 1·20 (0·95, 1·52) | 0·115 |
| | Strong res | 1·28 (1·03, 1·58) | **0·027** |
| | Mild res | 1·10 (0·93, 1·29) | 0·259 |
| | Low to no res | 1·08 (0·93, 1·26) | 0·296 |
| **Pandemic periods *w-1*** | | | 0·293[1] |
| | First wave | 1·21 (0·96, 1·53) | 0·100 |
| | Strong res | 1·23 (0·99, 1·53) | 0·062 |
| | Mild res | 1·10 (0·93, 1·29) | 0·257 |
| | Low to no res | 1·09 (0·94, 1·27) | 0·257 |
| **Pandemic periods *w-2*** | | | 0·190[1] |
| | First wave | 1·22 (0·97, 1·54) | 0·086 |
| | Strong res | 1·27 (1·02, 1·58) | **0·031** |
| | Mild res | 1·07 (0·91, 1·26) | 0·401 |
| | Low to no res | 1·10 (0·95, 1·28) | 0·207 |
| **COVID-19 intubation prevalence** | Week *w* | 1·04 (0·98, 1·09) | 0·197 |
| | Week *w-1* | 1·05 (0·99, 1·10) | 0·095 |
| | Week *w-2* | 1·05 (0·99, 1·11) | 0·078 |

[1]Global p-value

**Supplementary Table 9. Unadjusted estimates of incidence rate ratios (IRRs) associated to COVID-19-related variables for ESBL-producing *E. coli* in French hospitals and intensive care units (ICUs).** We depicted in bold the IRRs whose p-value is lower than 0·05.

| **ESBL-producing *E. coli*** | | **IRR** | **p-value** |
|---|---|---|---|
| **Hospital** | | | |
| **Pandemic periods *w*** | | | **<0·001**[1] |
| | First wave | 1·08 (1·02, 1·15) | **0·009** |
| | Strong res | 0·91 (0·85, 0·96) | **0·002** |
| | Mild res | 0·90 (0·87, 0·94) | **<0·001** |
| | Low to no res | 0·81 (0·79, 0·84) | **<0·001** |
| **Pandemic periods *w-1*** | | | **<0·001**[1] |
| | First wave | 1·07 (1·00, 1·13) | **0·039** |
| | Strong res | 0·89 (0·84, 0·95) | **<0·001** |
| | Mild res | 0·90 (0·87, 0·94) | **<0·001** |
| | Low to no res | 0·82 (0·79, 0·85) | **<0·001** |
| **Pandemic periods *w-2*** | | | **<0·001**[1] |
| | First wave | 1·07 (1·00, 1·14) | **0·036** |
| | Strong res | 0·88 (0·83, 0·94) | **<0·001** |
| | Mild res | 0·90 (0·87, 0·94) | **<0·001** |
| | Low to no res | 0·82 (0·79, 0·85) | **<0·001** |
| **COVID-19 intubation prevalence** | Week *w* | 1·00 (0·98, 1·02) | 0·949 |
| | Week *w-1* | 1·00 (0·98, 1·02) | 0·910 |
| | Week *w-2* | 0·99 (0·97, 1·01) | 0·455 |
| **ICU** | | | |
| **Pandemic periods *w*** | | | **<0·001**[1] |
| | First wave | 0·95 (0·78, 1·16) | 0·638 |
| | Strong res | 0·65 (0·53, 0·80) | **<0·001** |
| | Mild res | 0·86 (0·75, 0·99) | **0·037** |
| | Low to no res | 0·69 (0·61, 0·78) | **<0·001** |
| **Pandemic periods *w-1*** | | | **<0·001**[1] |
| | First wave | 1·04 (0·85, 1·26) | 0·720 |
| | Strong res | 0·65 (0·52, 0·80) | **<0·001** |
| | Mild res | 0·85 (0·74, 0·98) | **0·021** |
| | Low to no res | 0·71 (0·62, 0·81) | **<0·001** |
| **Pandemic periods *w-2*** | | | **<0·001**[1] |
| | First wave | 1·09 (0·89, 1·32) | 0·412 |
| | Strong res | 0·67 (0·55, 0·83) | **<0·001** |
| | Mild res | 0·80 (0·69, 0·92) | **0·001** |
| | Low to no res | 0·73 (0·64, 0·83) | **<0·001** |
| **COVID-19 intubation prevalence** | Week *w* | 0·93 (0·89, 0·98) | **0·013** |
| | Week *w-1* | 0·93 (0·88, 0·98) | **0·011** |
| | Week *w-2* | 0·92 (0·87, 0·97) | **0·003** |

[1]Global p-value

**Supplementary Table 10. Unadjusted estimates of incidence rate ratios (IRRs) associated to COVID-19-related variables for ESBL-producing *K. pneumoniae* in French hospitals and intensive care units (ICUs).** We depicted in bold the IRRs whose p-value is lower than 0·05.

| **ESBL-producing *K. pneumoniae*** | | **IRR** | **p-value** |
|---|---|---|---|
| **Hospital** | | | |
| **Pandemic periods *w*** | | | **0·005[1]** |
| | First wave | 1·19 (1·08, 1·31) | **<0·001** |
| | Strong res | 1·03 (0·93, 1·13) | 0·617 |
| | Mild res | 1·07 (1·00, 1·14) | **0·044** |
| | Low to no res | 1·02 (0·96, 1·08) | 0·560 |
| **Pandemic periods *w-1*** | | | **0·005[1]** |
| | First wave | 1·20 (1·09, 1·32) | **<0·001** |
| | Strong res | 1·02 (0·92, 1·12) | 0·728 |
| | Mild res | 1·06 (1·00, 1·13) | 0·058 |
| | Low to no res | 1·02 (0·96, 1·08) | 0·521 |
| **Pandemic periods *w-2*** | | | **0·022[1]** |
| | First wave | 1·17 (1·06, 1·29) | **0·001** |
| | Strong res | 1·02 (0·92, 1·13) | 0·694 |
| | Mild res | 1·06 (0·99, 1·13) | 0·099 |
| | Low to no res | 1·02 (0·96, 1·08) | 0·570 |
| **COVID-19 intubation prevalence** | Week *w* | 1·02 (1·00, 1·04) | 0·116 |
| | Week *w-1* | 1·02 (1·00, 1·04) | 0·120 |
| | Week *w-2* | 1·01 (0·99, 1·04) | 0·261 |
| **ICU** | | | |
| **Pandemic periods *w*** | | | **<0·001[1]** |
| | First wave | 1·33 (1·09, 1·62) | **0·005** |
| | Strong res | 0·77 (0·62, 0·95) | **0·016** |
| | Mild res | 1·19 (1·03, 1·36) | **0·017** |
| | Low to no res | 0·98 (0·86, 1·12) | 0·810 |
| **Pandemic periods *w-1*** | | | **<0·001[1]** |
| | First wave | 1·38 (1·14, 1·68) | **0·001** |
| | Strong res | 0·76 (0·61, 0·93) | **0·010** |
| | Mild res | 1·19 (1·03, 1·36) | **0·014** |
| | Low to no res | 0·98 (0·86, 1·11) | 0·736 |
| **Pandemic periods *w-2*** | | | **<0·001[1]** |
| | First wave | 1·44 (1·18, 1·74) | **<0·001** |
| | Strong res | 0·78 (0·63, 0·97) | **0·025** |
| | Mild res | 1·17 (1·02, 1·34) | **0·027** |
| | Low to no res | 1·00 (0·88, 1·13) | 0·944 |
| **COVID-19 intubation prevalence** | Week *w* | 1·00 (0·95, 1·05) | 0·909 |
| | Week *w-1* | 1·01 (0·96, 1·06) | 0·837 |
| | Week *w-2* | 1·01 (0·96, 1·06) | 0·727 |

[1]Global p-value

**Supplementary Table 11. Unadjusted estimates of incidence rate ratios (IRRs) associated to COVID-19-related variables for MRSA in French hospitals and intensive care units (ICUs).** We depicted in bold the IRRs whose p-value is lower than 0·05.

| **MRSA** | | **IRR** | **p-value** |
|---|---|---|---|
| **Hospital** | | | |
| **Pandemic periods *w*** | | | **<0·001[1]** |
| | First wave | 0·94 (0·87, 1·02) | 0·120 |
| | Strong res | 0·82 (0·76, 0·89) | **<0·001** |
| | Mild res | 0·84 (0·80, 0·88) | **<0·001** |
| | Low to no res | 0·75 (0·72, 0·79) | **<0·001** |
| **Pandemic periods *w-1*** | | | **<0·001[1]** |
| | First wave | 0·95 (0·88, 1·02) | 0·164 |
| | Strong res | 0·82 (0·76, 0·88) | **<0·001** |
| | Mild res | 0·83 (0·80, 0·88) | **<0·001** |
| | Low to no res | 0·75 (0·72, 0·79) | **<0·001** |
| **Pandemic periods *w-2*** | | | **<0·001[1]** |
| | First wave | 0·96 (0·89, 1·03) | 0·284 |
| | Strong res | 0·82 (0·76, 0·89) | **<0·001** |
| | Mild res | 0·84 (0·80, 0·88) | **<0·001** |
| | Low to no res | 0·75 (0·72, 0·79) | **<0·001** |
| **COVID-19 intubation prevalence** | Week *w* | 0·97 (0·94, 0·99) | **0·008** |
| | Week *w-1* | 0·97 (0·95, 1·00) | **0·027** |
| | Week *w-2* | 0·98 (0·95, 1·00) | **0·050** |
| **ICU** | | | |
| **Pandemic periods *w*** | | | **<0·001[1]** |
| | First wave | 0·64 (0·50, 0·82) | **<0·001** |
| | Strong res | 0·60 (0·47, 0·75) | **<0·001** |
| | Mild res | 0·64 (0·55, 0·75) | **<0·001** |
| | Low to no res | 0·65 (0·57, 0·75) | **<0·001** |
| **Pandemic periods *w-1*** | | | **<0·001[1]** |
| | First wave | 0·63 (0·49, 0·80) | **<0·001** |
| | Strong res | 0·59 (0·47, 0·75) | **<0·001** |
| | Mild res | 0·66 (0·57, 0·78) | **<0·001** |
| | Low to no res | 0·65 (0·56, 0·75) | **<0·001** |
| **Pandemic periods *w-2*** | | | **<0·001[1]** |
| | First wave | 0·64 (0·50, 0·81) | **<0·001** |
| | Strong res | 0·60 (0·47, 0·76) | **<0·001** |
| | Mild res | 0·66 (0·56, 0·78) | **<0·001** |
| | Low to no res | 0·65 (0·57, 0·76) | **<0·001** |
| **COVID-19 intubation prevalence** | Week *w* | 0·88 (0·83, 0·94) | **<0·001** |
| | Week *w-1* | 0·88 (0·83, 0·93) | **<0·001** |
| | Week *w-2* | 0·88 (0·82, 0·93) | **<0·001** |

[1]Global p-value

### d. Best model fits and estimates

**Supplementary Table 12. Estimates of the best count regression models of resistant infections in French hospitals and ICUs.** We depicted in bold the incidence rate ratios (IRRs) whose p-value is lower than 0·05.

| | | ESBL-producing *E. cloacae* complex | | ESBL-producing *K. pneumoniae* | | ESBL-producing *E. coli* | | MRSA | | CR *P. aeruginosa* | |
|---|---|---|---|---|---|---|---|---|---|---|---|
| | | **IRR (95% CI)** | **p** | **IRR (95% CI)** | **p** | **IRR (95% CI)** | **p** | **IRR (95% CI)** | **p** | **IRR (95% CI)** | **p** |
| **Hospital** | | | | | | | | | | | |
| Intercept | | **0·10 (0·10, 0·11)** | <0·001 | **0·21 (0·21, 0·21)** | <0·001 | **0·31 (0·30, 0·32)** | <0·001 | **0·22 (0·21, 0·22)** | <0·001 | **0·16 (0·15, 0·16)** | <0·001 |
| Incidence *w-1* | | **1·14 (1·11, 1·18)** | <0·001 | **1·08 (1·06, 1·10)** | <0·001 | **1·03 (1·01, 1·05)** | <0·001 | 0·99 (0·96, 1·01) | 0·306 | **1·07 (1·04, 1·09)** | <0·001 |
| Pandemic periods | First wave | - | - | - | - | 1·04 (0·97, 1·12) | 0·294 | 0·94 (0·87, 1·01) | 0·099 | - | - |
| | Strong | - | - | - | - | **0·91 (0·85, 0·98)** | 0·013 | **0·82 (0·76, 0·89)** | <0·001 | - | - |
| | Mild | - | - | - | - | **0·91 (0·86, 0·96)** | <0·001 | **0·84 (0·79, 0·88)** | <0·001 | - | - |
| | Low to none | - | - | - | - | **0·85 (0·81, 0·89)** | <0·001 | **0·74 (0·70, 0·78)** | <0·001 | - | - |
| COVID-19 intubation prev. *w-2* | | - | - | - | - | - | - | - | - | **1·06 (1·04, 1·09)** | <0·001 |
| Penicillins | | - | - | - | - | - | - | 1·01 (0·99, 1·03) | 0·266 | - | - |
| 3rd generation Cephalosporins | | 1·03 (1·00, 1·06) | 0·076 | **1·03 (1·01, 1·05)** | 0·007 | 1·01 (0·99, 1·03) | 0·325 | - | - | - | - |
| Imipenem + Meropenem | | - | - | - | - | - | - | - | - | **1·04 (1·01, 1·06)** | 0·003 |
| **ICU** | | | | | | | | | | | |
| Intercept | | **0·75 (0·71, 0·80)** | <0·001 | **1·14 (1·04, 1·25)** | 0·005 | **1·23 (1·12, 1·36)** | <0·001 | 0·91 (0·81, 1·03) | 0·142 | **2·01 (1·94, 2·08)** | <0·001 |
| Incidence *w-1* | | 1·04 (0·99, 1·11) | 0·14 | **1·07 (1·02, 1·13)** | 0·006 | 1·04 (0·98, 1·10) | 0·166 | 1·05 (0·98, 1·12) | 0·147 | **1·12 (1·08, 1·16)** | <0·001 |
| Pandemic periods | First wave | - | - | 1·18 (0·95, 1·48) | 0·129 | 0·92 (0·73, 1·16) | 0·486 | **0·67 (0·51, 0·87)** | 0·003 | - | - |
| | Strong | - | - | **0·76 (0·62, 0·94)** | 0·013 | **0·67 (0·54, 0·84)** | <0·001 | **0·63 (0·47, 0·82)** | <0·001 | - | - |
| | Mild | - | - | 1·11 (0·96, 1·28) | 0·146 | **0·86 (0·75, 1·00)** | 0·043 | **0·67 (0·54, 0·82)** | <0·001 | - | - |
| | Low to none | - | - | 1·03 (0·89, 1·18) | 0·708 | **0·73 (0·63, 0·85)** | <0·001 | **0·68 (0·58, 0·80)** | <0·001 | - | - |
| COVID-19 intubation prev. *w-2* | | - | - | - | - | - | - | - | - | **1·09 (1·05, 1·14)** | <0·001 |
| Penicillins | | - | - | - | - | - | - | 1·00 (0·92, 1·08) | 0·921 | - | - |
| 3rd generation Cephalosporins | | 1·05 (1·00, 1·12) | 0·064 | 1·05 (0·98, 1·13) | 0·141 | 1·02 (0·96, 1·10) | 0·491 | - | - | - | - |
| Imipenem + Meropenem | | - | - | - | - | - | - | - | - | 1·04 (0·99, 1·09) | 0·092 |

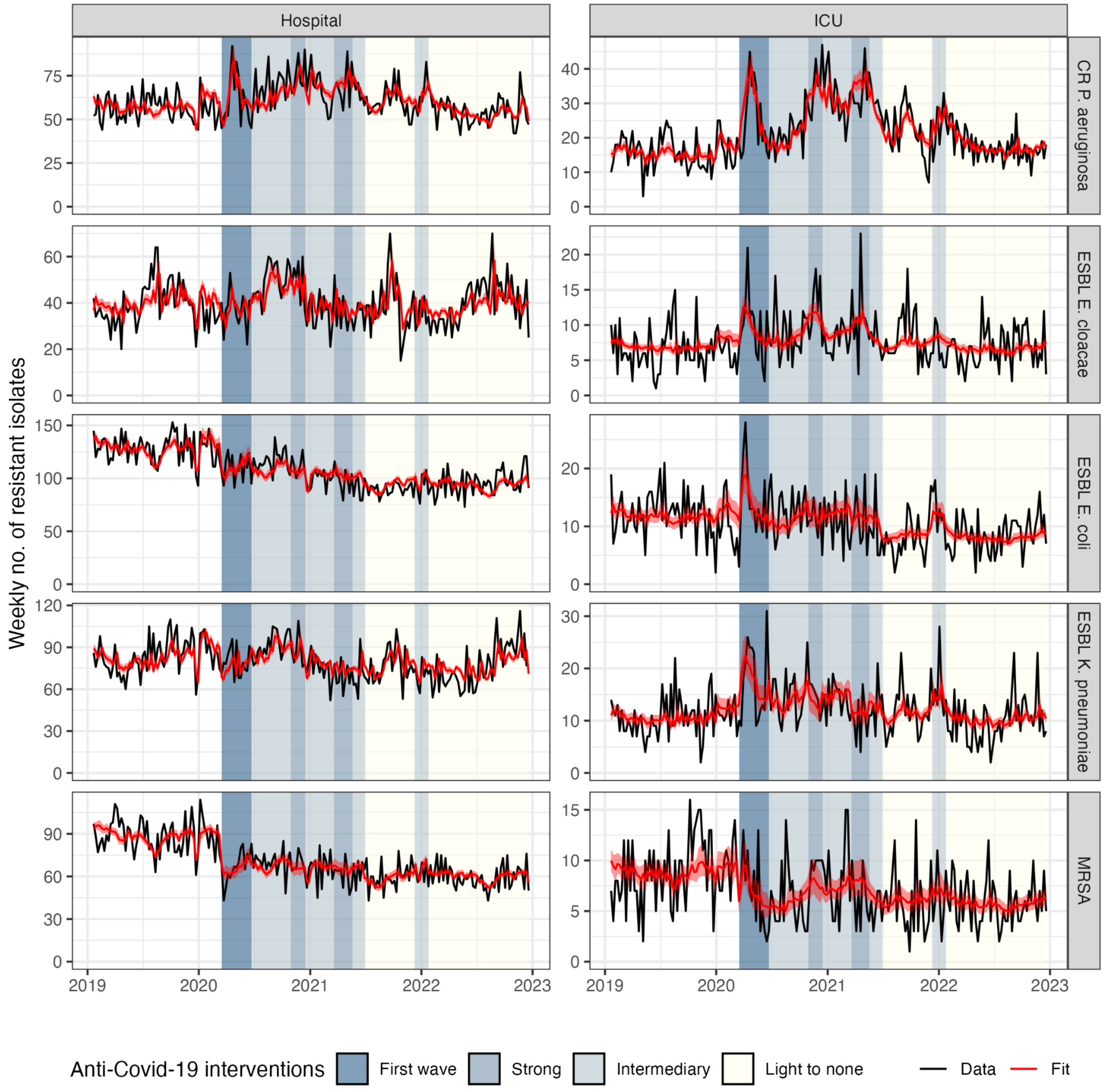


**Supplementary Figure 7. Fits of the best regression models applied to national incidence data.** The red ribbon indicates the 95% CI of the predicted weekly incidence of resistant bacteria.

### 6. Evolution in antibiotic use

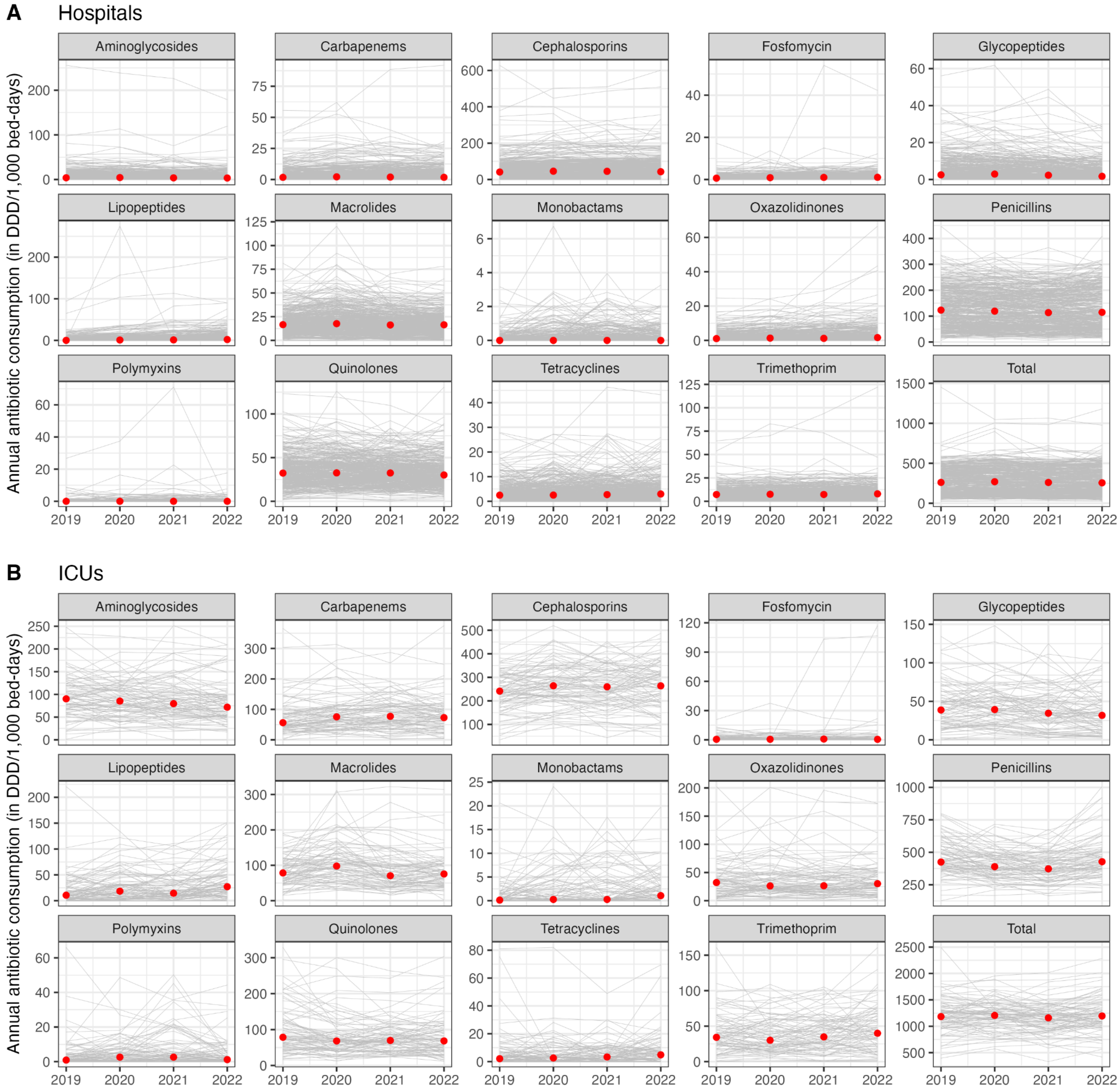


**Supplementary Figure 8. Annual antibiotic consumption by antibiotic class in (A) hospitals and (B) intensive care units (ICUs), 2019-2022.** Gray lines correspond to antibiotic consumption at the hospital or ICU level and red circles indicate the median consumption across all facilities. Consumption is expressed in defined daily doses (DDD) for 1,000 occupied bed-days.

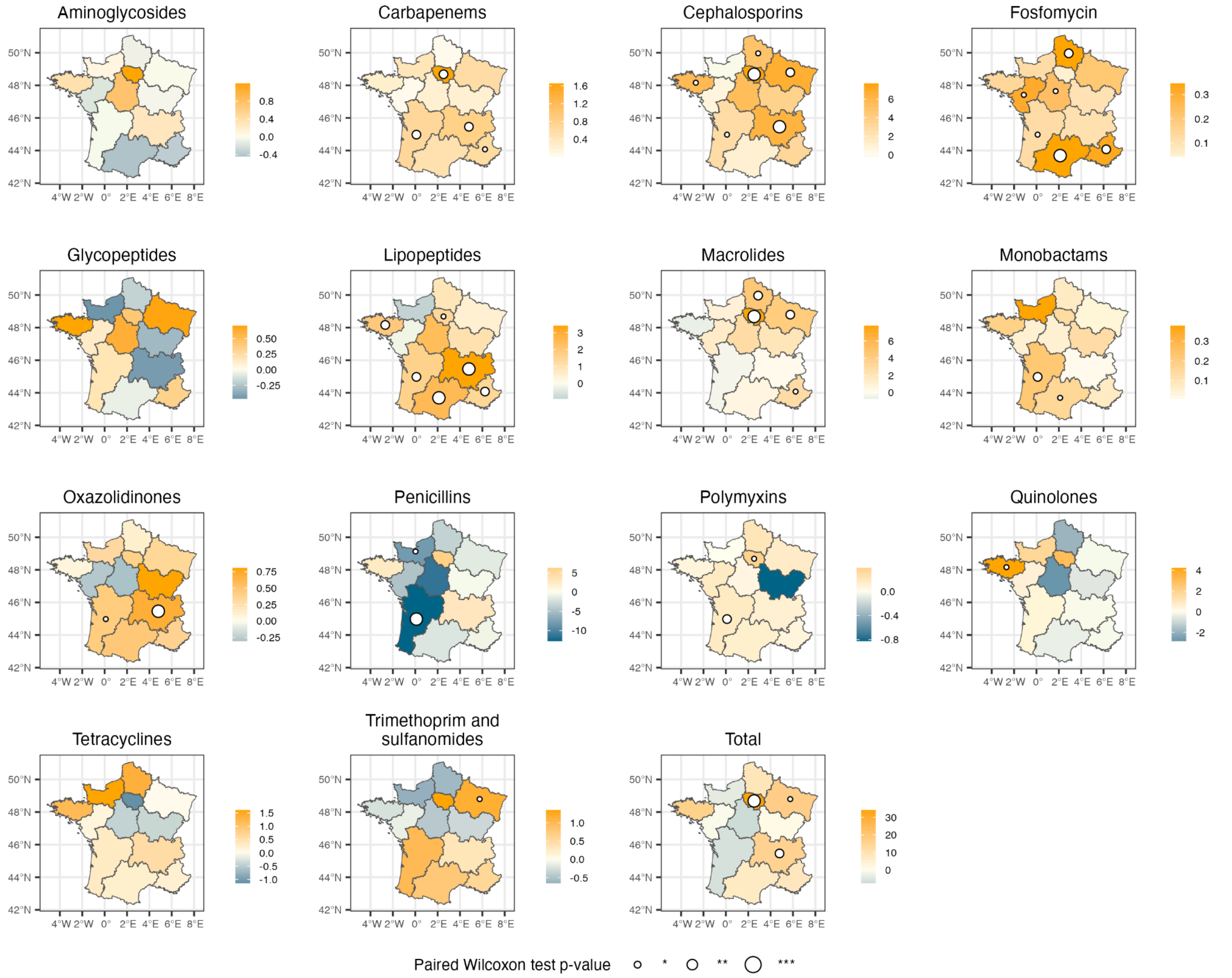


**Supplementary Figure 9. Median absolute change of antibiotic consumption in hospitals across regions between 2019 and 2020.** Positive values indicate an increase in 2020 and negative values a decrease. Each antibiotic class has its own absolute change gradient scale to ensure readability of absolute changes. We only report p-values≤0.05. *: p-value≤0.05; **: p-value≤0.01; ***: p-value≤0.001.

### 7. Sensitivity analysis exploring the impact of antibiotic use on incidence rate ratios estimates

In the principal multivariate analysis, we integrated the annual antibiotic use as a confusion factor. It has been shown that antibiotic consumption varied during the pandemic both in hospitals[7] and in the community.[8,9] This may have contributed to the modification of selective pressures in hospitals. In our dataset, we aggregated resistance data at the weekly level, however antibiotic use was only available annually and in hospitals. We evaluated whether not accounting for antibiotic use in hospitals changed our results.

For ESBL-KP, ESBL-EC, ESBL-ECC, and MRSA, we found the same best model as in the principal analysis (Supplementary Table 13) and incidence rate ratios (IRRs) barely changed (Supplementary Figure 10). This is indicative of little impact of target antibiotics used in hospitals for these four drug-bacterium pairs.

For CR-PA, we found the same model as in the principal analysis at the ICU level, but not at the hospital level (Supplementary Table 13). In the latter case, we tested both the model with pandemic periods *w-1* and the model with COVID-19 intubation prevalence *w-2* using a likelihood ratio test. The best model was the model with pandemic periods *w-1* ($p<1e-6$). Even though there is not a direct association with the prevalence of intubated COVID-19 patients, the incidence increased after the start of the first wave (Supplementary Figure 10) which is in agreement with the results of the principal analysis (Figure 4A).

**Supplementary Table 13. Results of model comparison when excluding antibiotic use.** Selected model is the one minimizing the AIC. We indicated in bold the AIC of the best selected model.

| | **CR *P. aeruginosa*** | **ESBL-producing *K. pneumoniae*** | **ESBL-producing *E. cloacae* complex** | **ESBL-producing *E. coli*** | **MRSA** |
|---|---|---|---|---|---|
| **Hospital** | | | | | |
| No COVID-19 variable | 1513 | **1607** | **1487** | 1628 | 1570 |
| Pandemic periods *w* | 1484 | 1608 | 1490 | **1584** | **1491** |
| Pandemic periods *w-1* | **1480** | 1610 | 1491 | 1591 | 1490 |
| Pandemic periods *w-2* | 1481 | 1612 | 1490 | 1591 | 1491 |
| COVID-19 intubation prevalence *w* | 1490 | 1608 | 1487 | 1630 | 1570 |
| COVID-19 intubation prevalence *w-1* | 1484 | 1608 | 1488 | 1629 | 1570 |
| COVID-19 intubation prevalence *w-2* | 1483 | 1609 | 1488 | 1628 | 1571 |
| **ICU** | | | | | |
| No COVID-19 variable | 1303 | 1190 | **1053** | 1164 | 1047 |
| Pandemic periods *w* | 1293 | **1181** | 1056 | **1146** | **1025** |
| Pandemic periods *w-1* | 1289 | 1179 | 1057 | 1146 | 1025 |
| Pandemic periods *w-2* | 1285 | 1182 | 1056 | 1147 | 1027 |
| COVID-19 intubation prevalence *w* | 1292 | 1192 | 1053 | 1162 | 1039 |
| COVID-19 intubation prevalence *w-1* | 1285 | 1192 | 1052 | 1162 | 1039 |
| COVID-19 intubation prevalence *w-2* | **1276** | 1192 | 1052 | 1160 | 1039 |

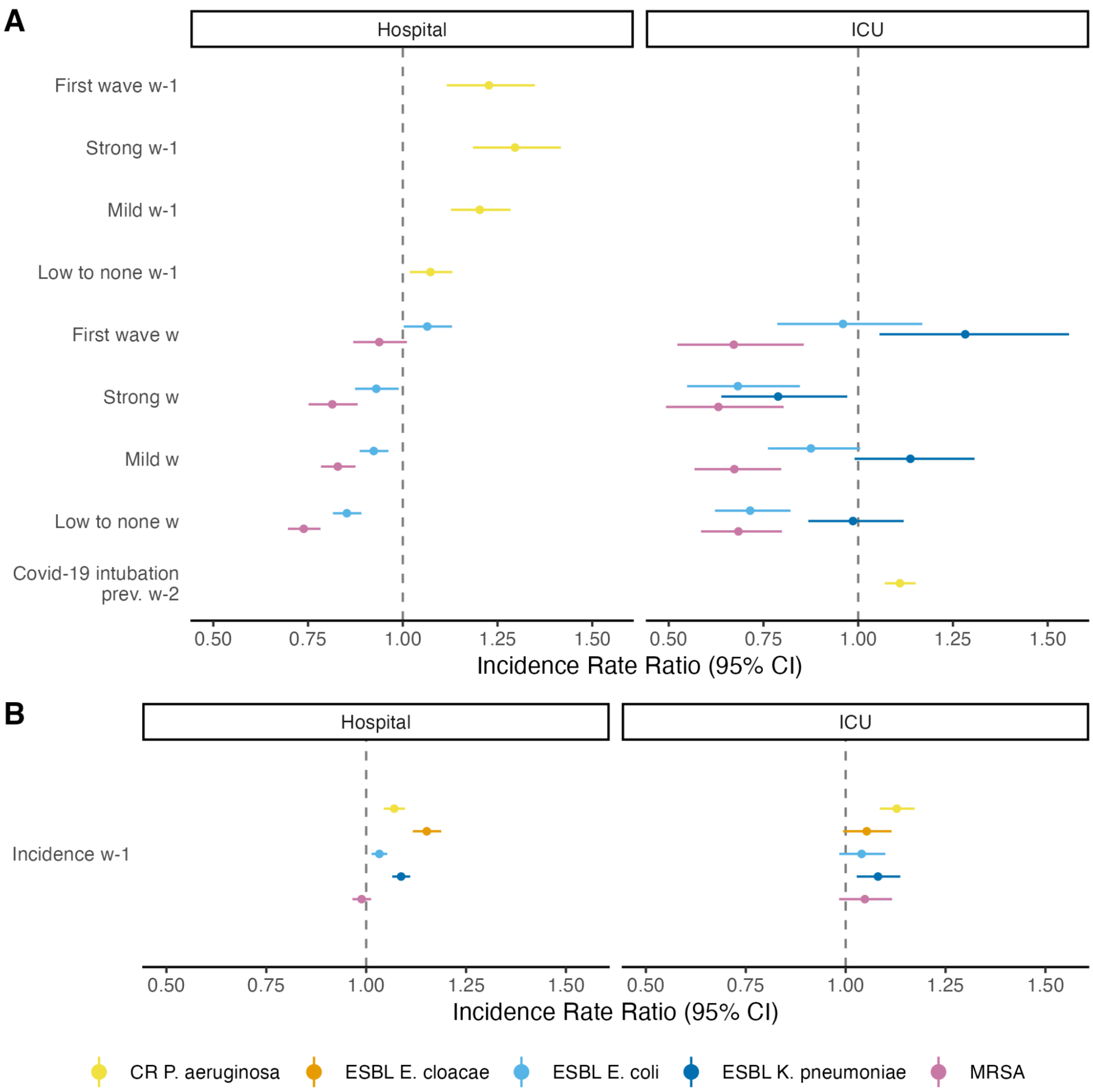


**Supplementary Figure 10. Results from the count regression analysis of resistant infections in French hospitals and intensive care units (ICUs).** (**A**) Incidence rate ratios (IRRs) of COVID-19-related variables for the best selected regression models. For ESBL-producing *E. cloacae* in ICUs and hospitals and *K. pneumoniae* in hospitals, the best models did not include COVID-19-related variables, thus they do not appear on the forest plots. For the other cases, the best models included either COVID-19-related periods at week *w*, COVID-19-related periods at week *w-1*, or the COVID-19 intubation prevalence at week *w-2*. IRR estimates for the COVID-19-related periods are relative to the pre-pandemic period. (**B**) Incidence rate ratios of the best regression models for the autocorrelation term.

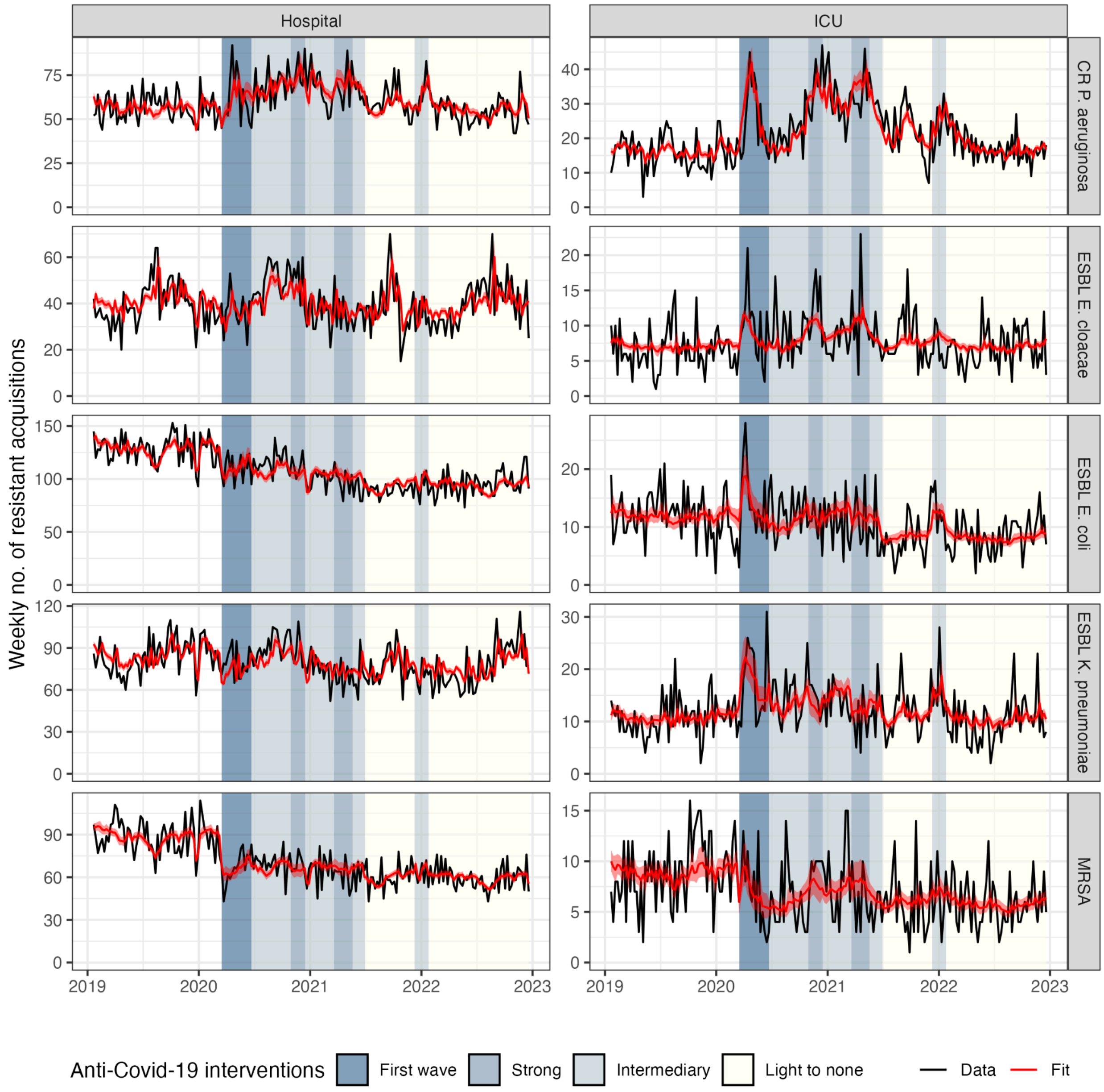


**Supplementary Figure 11. Fits of the best regression models applied to national incidence data in the sensitivity analysis without antibiotic consumption.** The red ribbon indicates the 95% CI of the predicted weekly incidence of resistant bacteria.

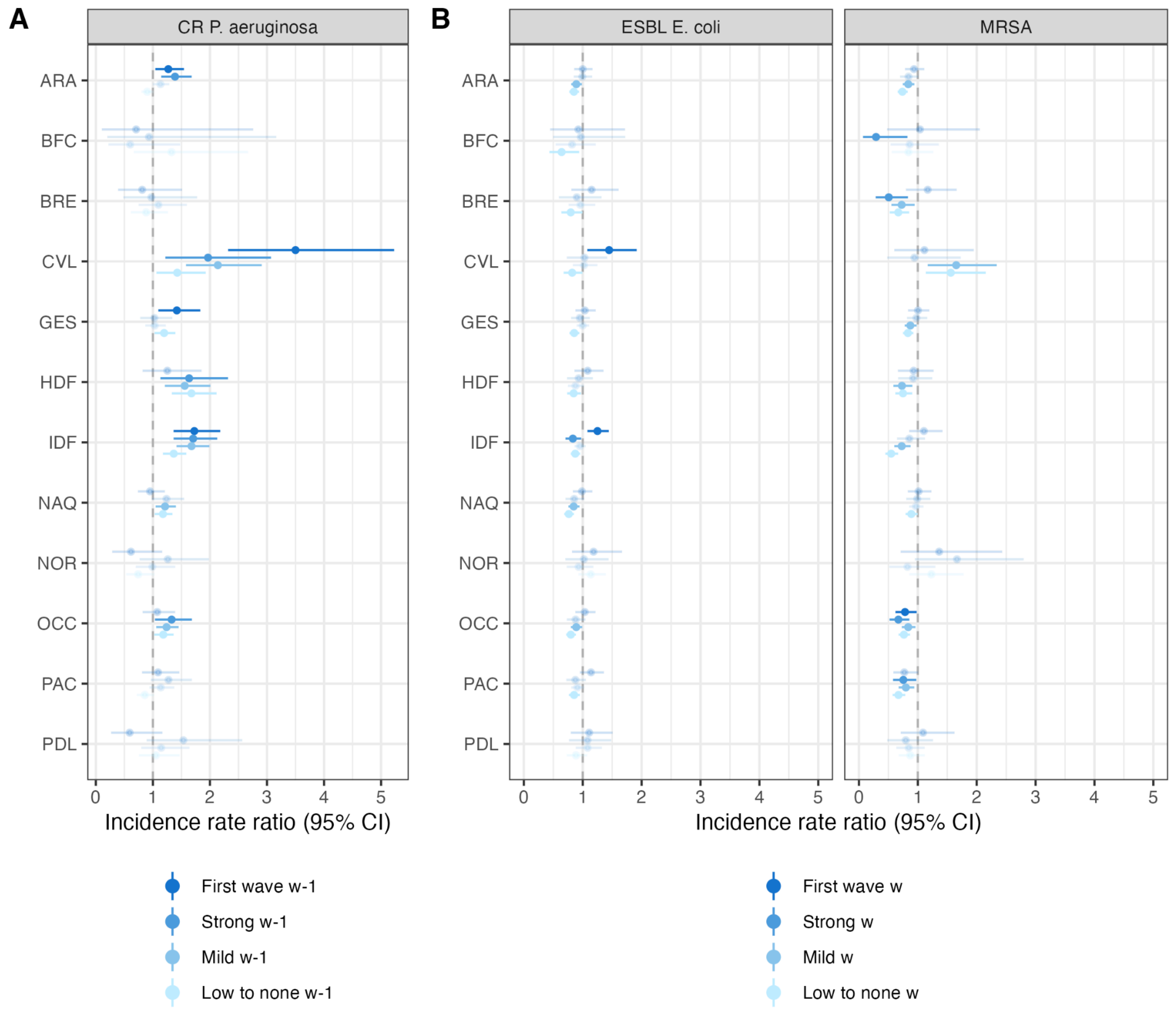


**Supplementary Figure 12. Regional heterogeneity of antibiotic resistance epidemiology during the pandemic in the sensitivity analysis without antibiotic use.** (**A**) Incidence rate ratios (IRRs) using the best model selected at the national level (without antibiotic consumption) on carbapenem-resistant *P. aeruginosa* (CR-PA) isolate incidence including the prevalence of COVID-19 intubated patients. (**B**) IRRs using the best model at the national level (without antibiotic consumption) on ESBL-producing *E. coli* (ESBL *E. coli*) and methicillin-resistant *S. aureus* (MRSA) isolate incidence including the pandemic periods at week *w*. Intervals correspond to the 95% CIs of the point estimates. Transparent IRRs have a p-value > 0·05. ARA: Auvergne-Rhône-Alpes; BFC: Bourgogne-Franche-Comté; BRE: Bretagne; CVL: Centre-Val de Loire; GES: Grand-Est; HDF: Hauts-de-France; IDF: Île-de-France ; NAQ: Nouvelle-Aquitaine; NOR: Normandie; OCC: Occitanie; PAC: Provence-Alpes-Côte d'Azur; PDL: Pays de la Loire.

8. **Sensitivity analyses for CR-PA isolates incidence**
   a. **Dynamics of CR-PA isolate incidence in hospitals is driven by intensive care units**

The results of the main analysis suggested that intubation of COVID-19 patients was associated with an increase in the incidence of CR-PA isolates in hospitals and ICUs at the national level. Given that intubation majorly concerns ICUs, we investigated whether the association between COVID-19 intubated patients and CR-PA isolate incidence was primarily driven by ICUs. To test this hypothesis, we applied the seven multivariate regression models to the incidence of CR-PA isolates in hospitals excluding isolates identified in ICUs. The best model did not include the COVID-19-related variables further suggesting that the increase in incidence was driven by patients hospitalized in ICUs (Supplementary Table 14).

**Supplementary Table 14. Akaike Information Criteria (AIC) of multivariate models applied to the incidence of CR-PA isolates.** The best model (in bold) is the one minimizing the AIC.

| | **CR *P. aeruginosa*** |
|---|---|
| **Hospital (except ICU)** | |
| No COVID-19 variable | **1356** |
| Pandemic periods *w* | 1358 |
| Pandemic periods *w-1* | 1360 |
| Pandemic periods *w-2* | 1360 |
| COVID-19 intubation prevalence *w* | 1358 |
| COVID-19 intubation prevalence *w-1* | 1358 |
| COVID-19 intubation prevalence *w-2* | 1358 |

   b. **Exploration of the causal relationship between COVID-19 related variables and CR-PA isolate incidence in hospitals and intensive care units**

To explore the robustness of our results and evidence for potential causality of the pandemic on CR-PA incidence, we tested models that integrate the prevalence of intubated COVID-19 patients in the following weeks (*w+1*, *w+2*, and *w+3*). We could show that the AIC of these models is as good as the AIC of the baseline model, without COVID-19 variables, and around 20 units higher than the AIC of the best model (Supplementary Figure 13A). Finally, we showed that the incidence peak during the first wave of the pandemic is better captured by the best model compared to the alternative models (Supplementary Figure 13C).

**A**

| Setting | Model | AIC |
|---|---|---|
| Hospital | Covid intubation w-2 (best model) | 1476 |
| Hospital | Baseline model | 1496 |
| Hospital | Baseline model | 1356 |
| Hospital | Covid intubation w+1 | 1494 |
| Hospital | Covid intubation w+2 | 1501 |
| Hospital | Covid intubation w+3 | 1505 |
| ICU | Covid intubation w-2 (best model) | 1275 |
| ICU | Baseline model | 1291 |
| ICU | Covid intubation w+1 | 1292 |
| ICU | Covid intubation w+2 | 1293 |
| ICU | Covid intubation w+3 | 1294 |

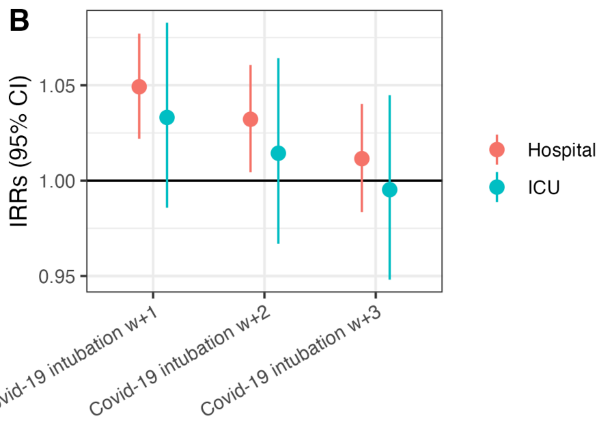


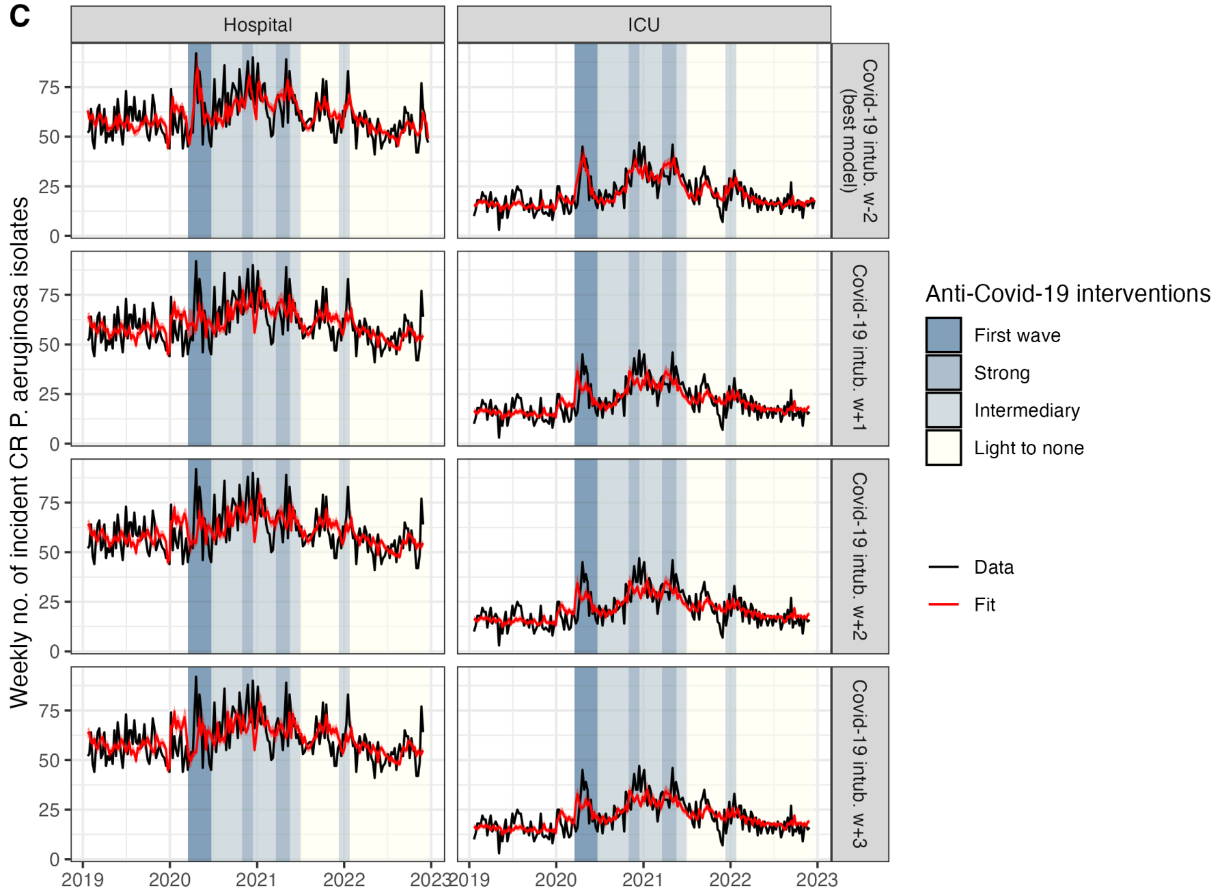


**Supplementary Figure 13. Investigations of the temporal associations between CR-PA and COVID-19 intubated patients.** (**A**) AIC of the best model selected in the national analysis that includes the prevalence of COVID-19 patients at week *w-2*, and of two alternative models that integrate the prevalence of intubated COVID-19 patients at week *w+1* or *w+2*. At both the hospital and ICU levels, the model with COVID-19 data from the preceding week best explains the data. (**B**) Incidence rate ratios and their 95% CI for the models with COVID-19 intubation at weeks *w+1* or *w+2* at the hospital and ICU levels. (**C**) Fits of the best model (COVID-19 intubation w-2) and the alternative models at the hospital and ICU levels. Data in black corresponds to the weekly number of incident CR-PA. Fits are represented in red with the red ribbon corresponding to the 95% CI.
